# Exploring Electroencephalography for Chronic Pain Biomarkers: A Large-Scale Benchmark of Data- and Hypothesis-Driven Models

**DOI:** 10.64898/2026.03.06.26347785

**Authors:** Felix S. Bott, Özgün Turgut, Paul Theo Zebhauser, Divya B. Adhia, Yoni K. Ashar, Melissa A. Day, Yelena Granovsky, Mark P. Jensen, Tor D. Wager, David Yarnitsky, Daniel Rueckert, Markus Ploner

**Author notes:** joint first authors (Felix S. Bott, Özgün Turgut).

## Abstract

Resting-state electroencephalography (EEG) has been proposed as a scalable source of biomarkers for chronic pain, but its clinical potential remains uncertain. To systematically evaluate this potential, we benchmarked nine modeling strategies, spanning conventional machine learning with handcrafted features to state-of-the-art deep learning. Across 72 configurations of signal representations and model architectures, we trained models to predict self-reported pain intensity, using chronological age decoding as a positive control. Pain prediction performance was limited (R=0.15), with the best results achieved by conventional connectivity-based models. In contrast, age was robustly decoded from the same dataset (R=0.53), confirming technical efficacy. These findings indicate that resting-state EEG contains limited information about inter-individual differences in chronic pain intensity, making it unlikely to yield clinically actionable biomarkers in cross-sectional settings. Instead, its potential may lie in intra-individual modeling of pain dynamics, which could advance individualized mechanistic insights and more personalized treatment of chronic pain.

## Introduction

Developing neuroimaging-based biomarkers to diagnose, monitor, and predict chronic pain is a priority in translational pain research [1–7]. Electroencephalography (EEG), in particular, has attracted considerable interest as a scalable, non-invasive, and widely accessible tool. However, despite increasing methodological sophistication, EEG-based signatures of chronic pain have yielded modest effect sizes and limited generalizability, particularly regarding inter-individual differences in pain intensity [1, 5, 8–10]. These limitations raise a fundamental question: do current results reflect intrinsic biological constraints on the information content of resting-state EEG, or are they consequences of suboptimal modeling strategies?

Recent efforts partially addressed this question by curating and analyzing a large, multi-site dataset comprising EEG recordings collected by research groups in Australia, Germany, Israel, New Zealand, and the United States, recorded under heterogeneous conditions [1]. With EEG recordings from more than 600 individuals with chronic pain, this constituted the largest analysis of EEG biomarkers in this population to date. In that work, we applied a strongly *hypothesis-driven* modeling strategy focusing on connectivity between large-scale functional brain networks [11]. While strong evidence for associations between EEG-derived functional connectivity patterns and chronic pain intensity was found under these favorable conditions of large sample size and biologically motivated a priori feature selection, the modeling strategies showed limited generalizability and explained a low proportion of variance in pain ratings. It remains unclear whether these limitations were driven by intrinsic signal constraints or could be mitigated by relaxing a priori assumptions, employing purely *data-driven* modeling strategies such as state-of-the-art deep learning.

In this work, we addressed this uncertainty by reanalyzing the EEG recordings of 623 individuals with chronic pain using a comprehensive range of data-driven modeling strategies. We evaluated their effectiveness in extracting neural signatures from EEG that can serve as potential biomarkers of chronic pain severity. Our study spans both conventional machine learning relying on handcrafted, *a priori defined* signal features, and state-of-the-art deep learning (DL) models that *learn* features directly from the signal. Within the DL domain, we investigated both convolutional neural networks, trained directly on the task, and Transformers [12]. The Transformer models were pre-trained in a self-supervised manner on large-scale, unlabeled EEG data before being fine-tuned on the specific target task. This pre-training paradigm enables the learning of general signal features from large amounts of task-agnostic data, capturing information that fully supervised learning might miss due to the lack of task-specific data. In total, we systematically evaluated 72 distinct configurations of signal representations and model architectures to assess the efficacy of data-driven modeling strategies for EEG-based biomarker discovery. This approach aims to systematically characterize the feasibility, limitations, and translational potential of EEG-based biomarkers for inter-individual differences in chronic pain.

Crucially, to validate the technical sensitivity of our modeling strategies to extract biological information from EEG, we introduced chronological age prediction as a positive control task. Given the well-established associations between age and EEG signals [13], successful age decoding serves as a necessary technical validation of modeling efficacy. This establishes a performance baseline within the same harmonized dataset, ensuring the subsequent findings on chronic pain prediction can be confidently attributed to the intrinsic information content of the EEG signal, unconfounded by constraints of the modeling strategies. Ultimately, by systematically benchmarking diverse modeling strategies, our study aims to determine whether powerful data-driven approaches such as deep learning can overcome the current bottleneck in chronic pain biomarker development. An overview of our study and the research questions is provided in Figure 1.

The study and its analysis were preregistered at the OSF Registries (https://osf.io/9j3xh/).

**Figure 1:**
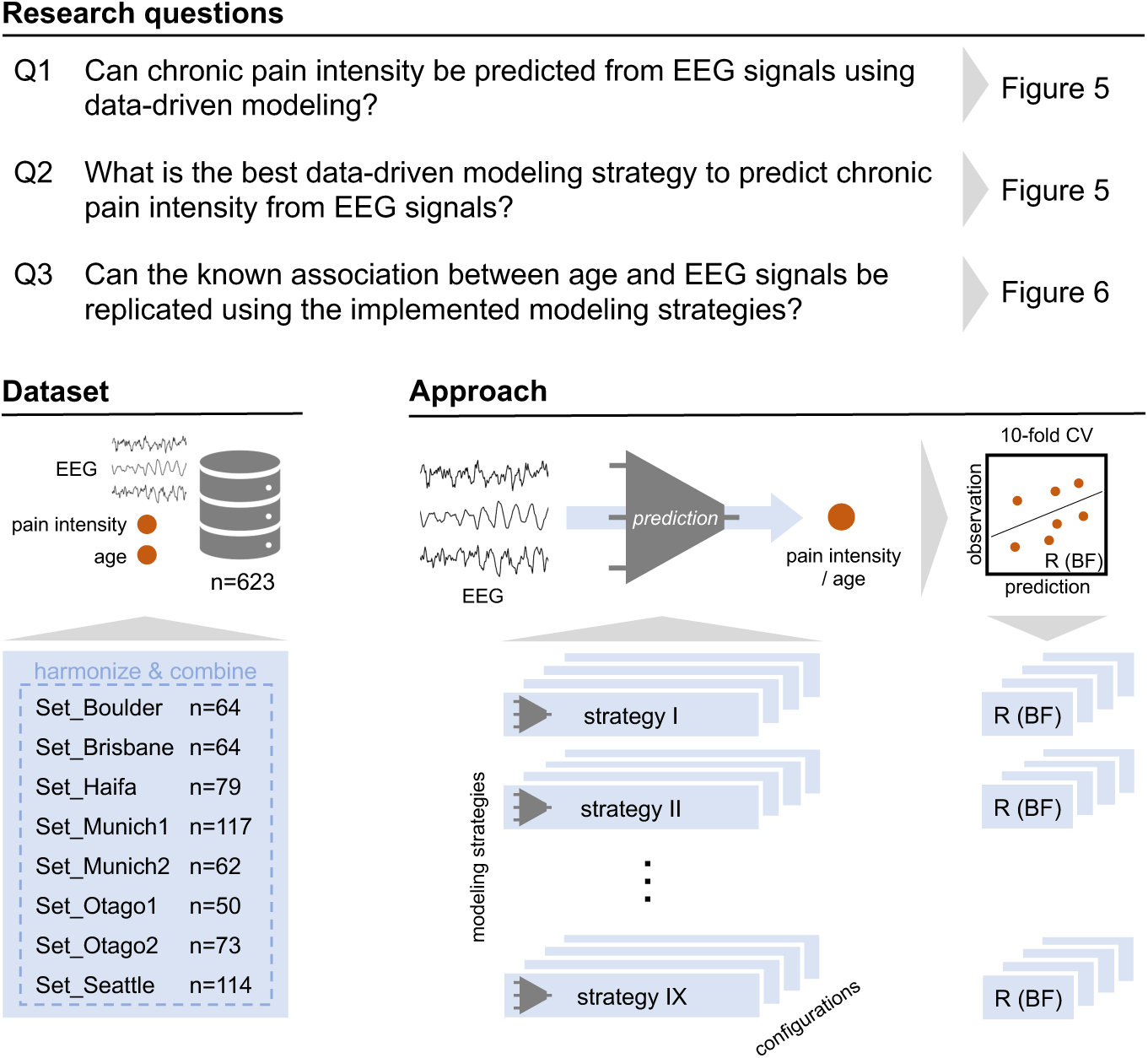
Study overview. Summary of research questions, dataset and analysis approach.

## Results

### Overview of modeling strategies

To comprehensively evaluate the potential of EEG for chronic pain biomarker discovery, we designed a large-scale benchmarking study spanning distinct hypothesis-driven and data-driven modeling strategies, as summarized in Figure 2. Models were trained and evaluated on a multi-site dataset [1] comprising resting-state EEG recordings, self-reported pain intensity, and demographic information from 623 individuals with chronic pain. Our evaluation strategy addressed two fundamental questions: first, whether data-driven deep learning can extract neural signatures of chronic pain that hypothesis-driven machine learning misses; and second, whether findings on chronic pain biomarkers can be confidently attributed to the intrinsic information content of the EEG signal, free of technical bias and modeling constraints. To address the latter, we introduced chronological age prediction as a positive control task. Given that age has been robustly linked to EEG [13], successful age decoding serves as a necessary technical validation of the modeling strategies. Model performance was quantified using 10-fold cross-validated Pearson correlation coefficients (R) between predicted and observed values. To determine statistical evidence for or against the presence of predictive structures, we computed Bayes Factors (BF), interpreting BF > 3 and BF > 10 as moderate and strong evidence for an effect, respectively, and BF < 1/3 as evidence against an effect. We selected prediction-observation correlation as the primary performance metric to reflect our intention to test for an in-principle association between EEG signals and pain intensity, rather than to evaluate absolute predictive accuracy (e.g., R^2^), which we report for completeness [14].

**Figure 2:**
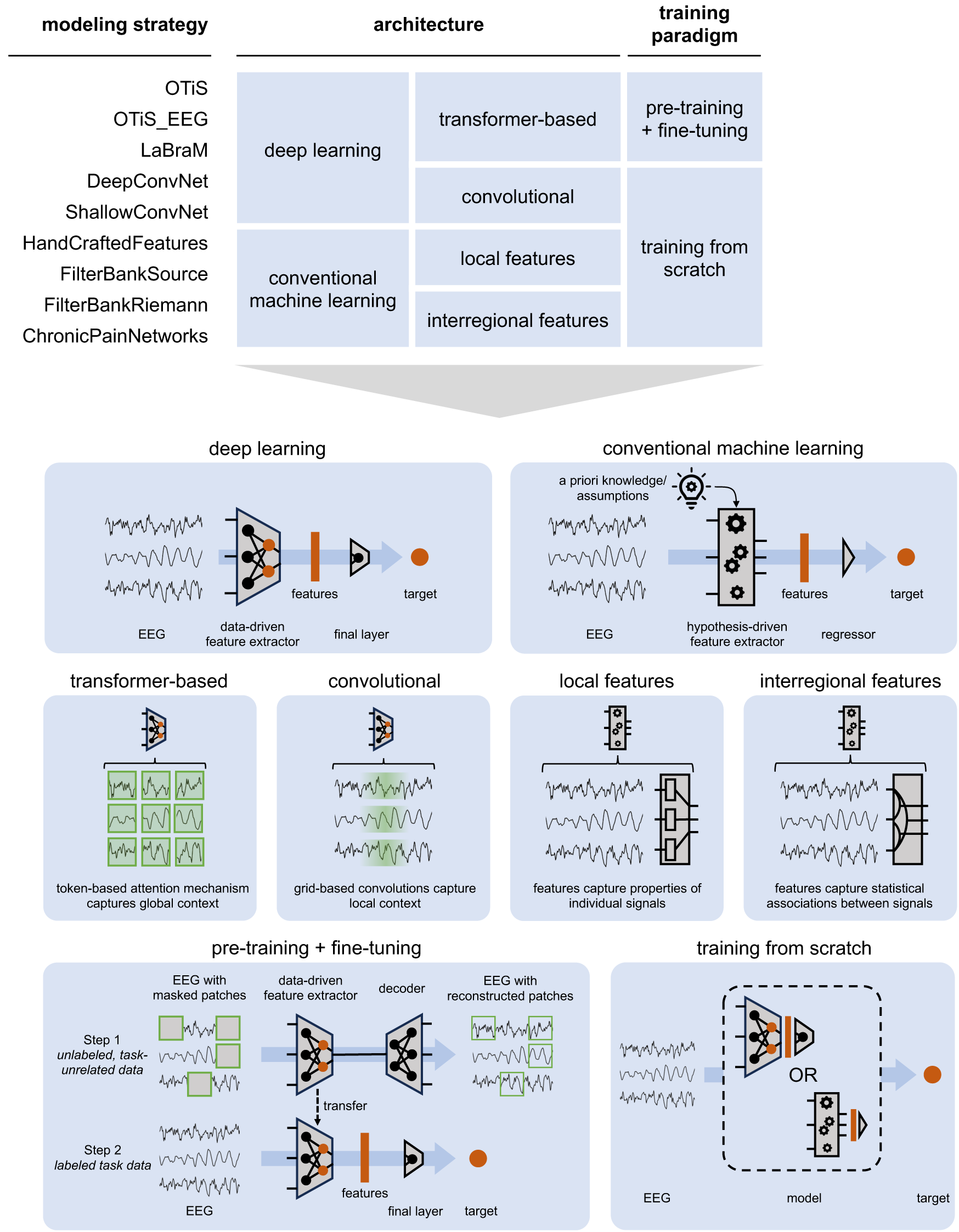
Overview of modeling strategies. Top: List of modeling strategies and corresponding categorizations of model architectures and training paradigms. Bottom: Explanatory illustrations of model architectures and training paradigms.

We first explored state-of-the-art deep learning, specifically focusing on the trade-off between general-purpose and domain-specific pre-training in Transformer architectures [12]. To test whether exposure to diverse signals facilitates the learning of powerful features applicable to EEG, we deployed *OTiS* [15], a state-of-the-art general-purpose model pre-trained on a large corpus including ECG, EEG, audio, and finance signals. This was contrasted with *LaBraM* [16], a specialized model exclusively designed and trained for EEG signals, to assess whether domain-specific pre-training yields superior features. To rigorously isolate the effects of domain specialization from the influence of architectural differences and training strategies, we further included variants of *OTiS_EEG* [17]. This control model shares the identical architecture and training strategy with *OTiS* but was pre-trained solely on the EEG subset of the original pre-training corpus. This design enables a direct evaluation of whether the inclusion of signals from other domains enhances feature learning for EEG analysis compared to pre-training on EEG signals alone. Complementing the Transformer approaches, we evaluated established convolutional neural networks (ConvNets) to assess deep learning performance on EEG in a fully supervised setting, avoiding the complexity of self-supervised pre-training. This included *DeepConvNet* [18], a deep architecture comprising four convolutional blocks, and *ShallowConvNet* [18], a lightweight architecture inspired by filter-bank common spatial patterns successfully applied in brain-computer interfaces [19]. These models are widely established in EEG analysis and serve as a baseline for the more complex Transformer architectures.

Finally, to benchmark these deep learning strategies against conventional machine learning, we evaluated models relying on handcrafted, a priori defined features. We examined modeling strategies focusing on local signal features, including *HandCraftedFeatures* [20], which utilizes simple sensor-level signal statistics in the time and frequency domain, and *FilterBankSource* [13, 21], which relies on spectral power of source-reconstructed signals in different frequency bands and cortical regions. These were compared against modeling strategies capturing interregional signal features, i.e., functional connectivity, such as *FilterBankRiemann* [22, 23], which utilizes features from covariance matrices of sensor-level signals in distinct frequency bands, and *ChronicPainNetworks* [1], which utilizes amplitude-based connectivity within seven functional brain networks. This hierarchical and extensive comparison, ranging from general-purpose models to specific handcrafted features, allowed us to systematically probe the nature of pain-related information in EEG signals.

### Configuration of modeling strategies

While the approaches defined above represent high-level conceptual modeling strategies, their success depends on specific implementation choices regarding signal representation and model architecture. To disentangle these factors, we systematically explored a design space of 72 distinct configurations, as summarized in Figure 3. This analysis was needed to isolate the optimal conditions for EEG-based biomarker discovery; first, by evaluating how EEG signals are best *presented* to the models, and second, by evaluating how models optimally *transform* these signals into accurate clinical predictions. These configurations were validated for predicting both chronic pain intensity and chronological age, as detailed in Supplementary Figures 1 and 2.

**Figure 3:**
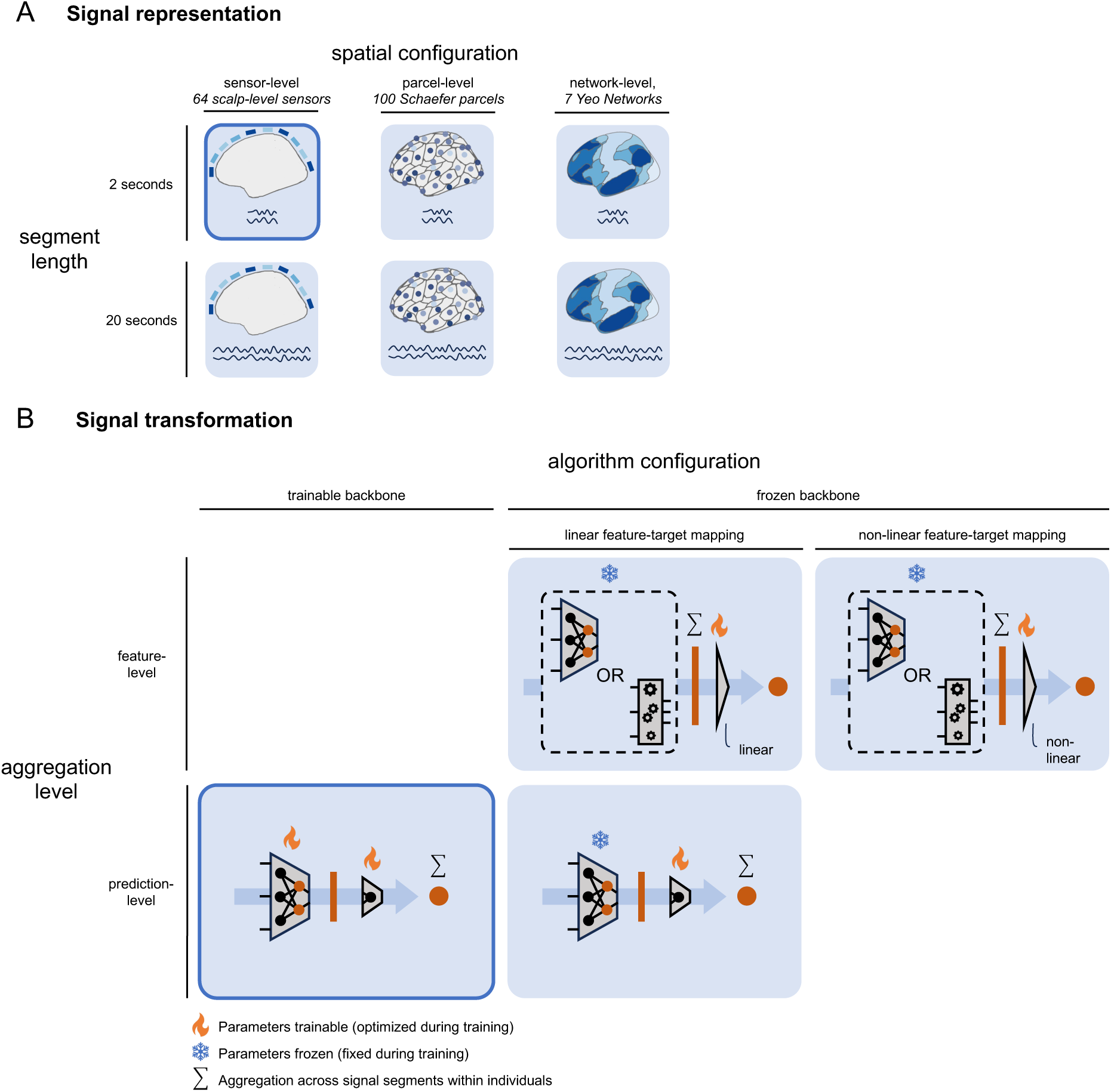
Configuration of modeling strategies. **(A)** Input signal representations varied along spatial and temporal dimensions. Spatially, input signals represented either the default sensor-level (64 standard-layout EEG channels), parcel-level (sensor-level signals source-projected to centroids of 100 parcels of Schaefer-atlas atlas), or network-level (parcel-level signals aggregated within 7 functional networks defined by the Yeo atlas). Temporally, input signals were segmented into durations of either the default 2 seconds or 20 seconds. **(B)** Model architecture configurations varied in feature extraction and prediction mechanisms. To extract features, models utilized either a default trainable backbone or a frozen backbone. To map features to clinical targets, we compared feature-level aggregation (averaging features across segments within individuals before prediction) against the default prediction-level aggregation (averaging predictions across segments within individuals). Finally, the prediction head employed either a default linear or a non-linear mapping. **Blue** highlighting indicates the default configuration, selected to serve as a robust, assumption-free baseline.

#### Signal Representation: Spatial and Temporal Configuration

To determine how EEG signals are best presented to the model, we varied the input signal along spatial and temporal dimensions. *Spatially*, we compared three levels of abstraction: the default sensor-level (64 scalp-level sensors), representing the raw scalp signal; parcel-level (100 Schaefer parcels), representing source-projected brain activity within 100 parcels of the Schaefer atlas to assess the benefit of anatomically informed signal unmixing; and network-level (7 Yeo networks), obtained by aggregating parcel-level signals within large-scale functional brain networks to assess the benefit of functionally-informed dimensionality reduction. Given the long duration of resting-state EEG signals, they are typically split into temporal segments that are processed independently by the model. Hence, *temporally*, we varied the input signal length between 2 seconds and 20 seconds. Default 2-second signal segments capture rapid neural dynamics, while with 20-second segments we test the hypothesis that pain intensity may be encoded in slower, accumulating signal patterns ranging over longer periods.

#### Signal Transformation: Feature Extraction and Prediction

Once the signal is presented to the model, the challenge shifts to how the model transforms this input into a clinical prediction. We analyzed this process across two distinct stages: the extraction of features from the input, and the subsequent mapping of those features to the target. First, to determine the optimal *feature extraction* mechanism, we compared linear probing (frozen backbone; the feature-extractor remains fixed during training) against full fine-tuning (trainable backbone; the feature extractor is optimized during training). Because linear probing crucially depends on the informative value of the pretrained representations, strong performance in this setting indicates that these features already capture clinically relevant signal. By evaluating both linear probing and full fine-tuning, we assess the trade-off between the greater training stability and lower overfitting risk of linear probing and the potential gains from task-specific adaptation through full fine-tuning. Second, we examined the optimal mapping of these features to targets. To determine the most effective *aggregation strategy* to reintegrate information of the segmented EEG signal within individuals, we compared feature-level aggregation (averaging features across segments within individuals before prediction) and prediction-level aggregation (averaging predictions across segments within individuals). This comparison reveals whether accurate predictions emerge from feature stability across signal segments or from the consensus of independent segment predictions. Simultaneously, we determined the optimal *prediction head complexity* by comparing linear and non-linear heads, testing whether the relationship between features and clinical targets is direct or requires modeling complex non-linear relationships.

Based on these considerations, we established a default configuration for each modeling strategy to serve as a simple and robust baseline. For the input representation, we selected sensor-level (64 channel) signals and utilized 2-second segments. For deep learning models, the default signal transformation consisted of a trainable backbone combined with prediction-level aggregation across segments. For models with a frozen backbone, we used linear mappings from features to targets as the default prediction head.

### Dataset harmonization

Given the multi-site nature of the curated dataset, as depicted in Figure 4, reducing heterogeneity across resting-state EEG signals from 623 individuals was essential to mitigate potential confounds arising from different acquisition systems and conditions. Consequently, prior to modeling, we performed rigorous dataset harmonization to ensure that detected patterns reflected genuine neural signatures rather than site-related artifacts. EEG signals were rescaled within each dataset such that their average logarithmic global power was equalized. Crucially, we adopted dataset-wise standardization rather than subject- and channel-wise standardization. This was critical to preserve inter-individual differences in absolute signal power, a feature hypothesized to carry pain-relevant information [24]. We further standardized the clinical targets, including pain intensity and chronological age, by z-scoring them within each dataset. This prevented models from exploiting trivial associations between site-specific demographics and acquisition conditions, thereby forcing the models to learn robust cross-site biomarkers.

**Figure 4:**
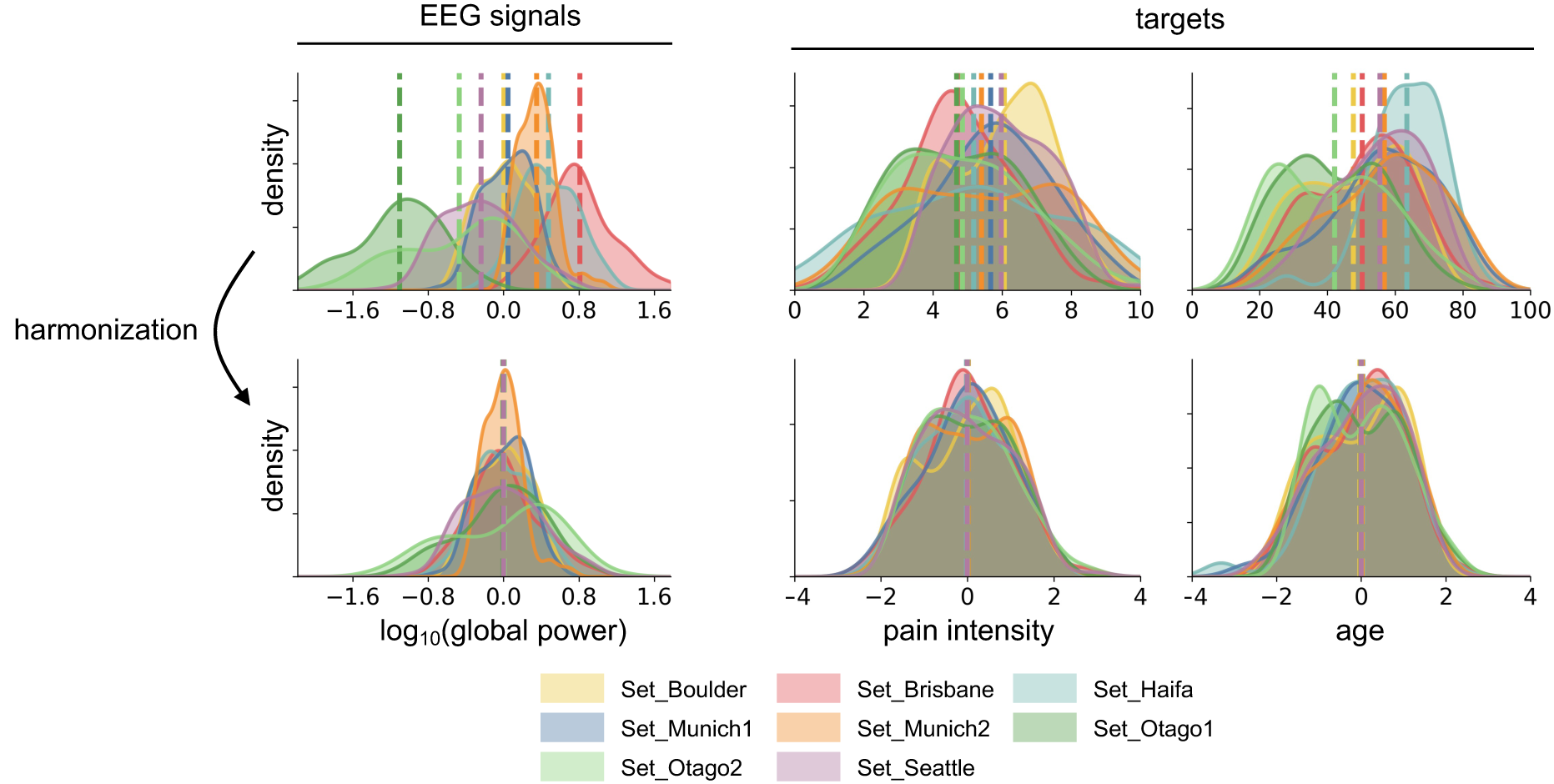
Cross-site dataset harmonization. **(Left)** EEG Signal Harmonization. *Top:* Distributions of logarithmic global power across individuals for each constituent dataset prior to harmonization, revealing systematic site-specific offsets due to varying recording equipment and conditions. *Bottom:* Corresponding distributions after dataset-wise rescaling. This step equalized the average global power across datasets to mitigate site artifacts, while crucially preserving potentially informative inter-individual differences in absolute power within each dataset. **(Right)** Target Harmonization. *Top:* Distributions of chronic pain intensity ratings and age prior to harmonization, indicating some clinical and demographic heterogeneity across sites. *Bottom:* Corresponding distributions after dataset-wise z-scoring. Harmonizing both signals and targets prevents models from exploiting trivial site-level differences for prediction.

### Pain prediction benchmark

Despite the extensive exploration of 72 modeling configurations, the prediction of chronic pain intensity from resting-state EEG remained limited, as summarized in Figure 5. Overall, the modeling strategies yielded low predictive performance, with cross-validated correlations between predicted and observed pain intensity ratings ranging between R=-0.02 and R=0.15. The proportion of variance explained in pain intensity ratings was consistently below R^2^=0.01. Importantly, rank-based prediction-observation correlations (Spearman’s rho) aligned closely with the parametric measures (Pearson’s R), arguing against outlier-driven effects and confirming the stability of these low correlations.

**Figure 5:**
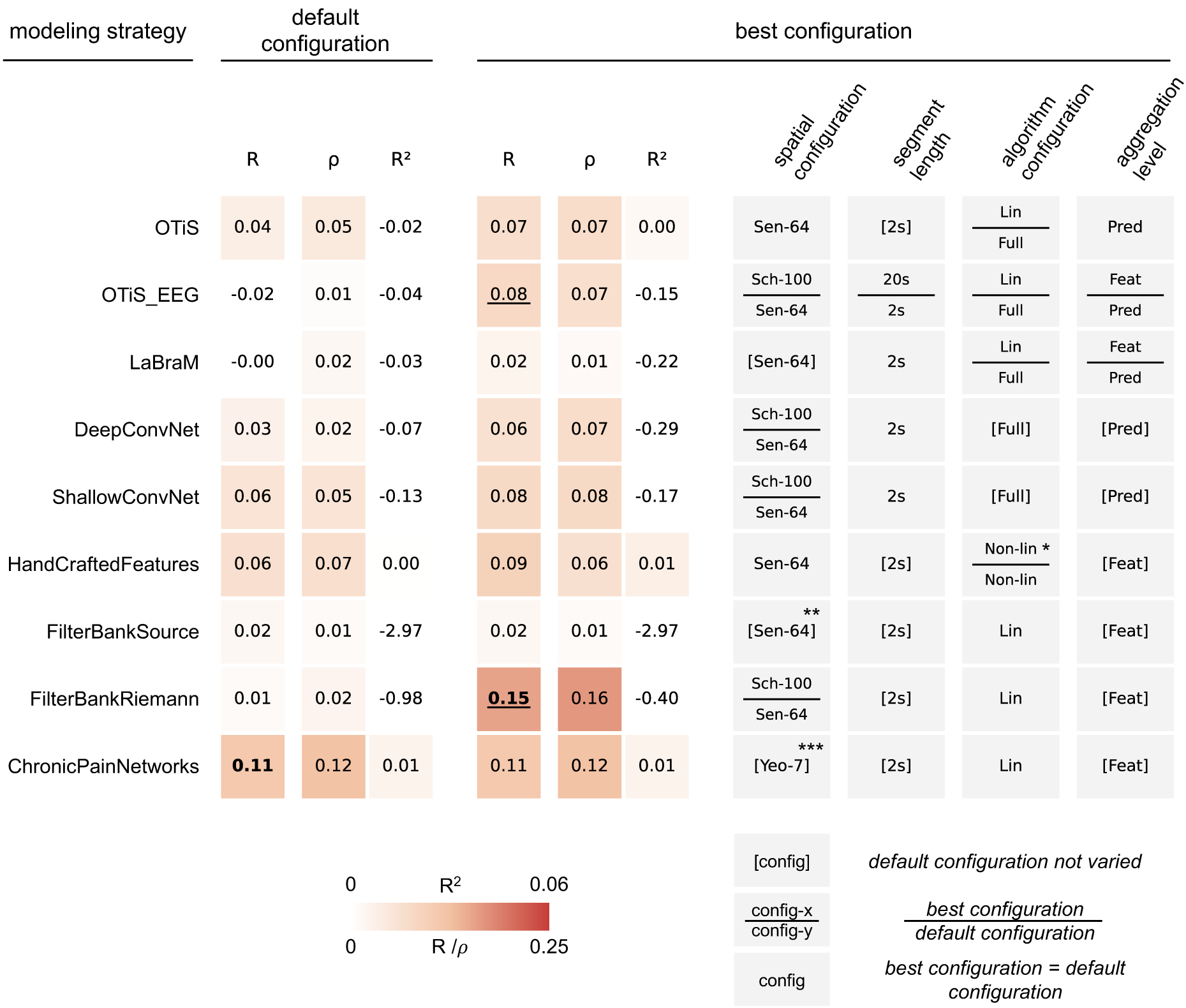
Pain intensity prediction performance of different modeling strategies. Cross-validated measures of association between predicted and observed pain intensity are reported (Pearson’s R; Spearman’s ρ; coefficient of determination R^2^). Scores in **bold** indicate at least moderate evidence for performance above chance (BF > 3). **(Left)** Default Configuration. This block summarizes the performance of each modeling strategy in its default configuration (see Figure 3). **(Right)** Best Configuration. This block summarizes the performance of the single best configuration found for each modeling strategy, alongside its specific configuration details. <u>Underlined</u> scores indicate a BF ratio of best configuration versus default larger than 10. Brackets [] indicate configurations which were not varied for a given modeling strategy. Tiles divided by a horizontal bar indicate configurations for which the best configuration (top entry) differed from the default configuration (bottom entry). Sen-64: 64 sensor-level signals, Sch-100: 100 parcel-level signals, Yeo-7: 7 network-level signals, Lin: linear mapping from features to targets, Non-lin: non-linear mapping from features to targets, Full: full network training, Pred: prediction-level aggregation, Feat: feature-level aggregation. *The default configuration employed a non-linear mapping using a random forest algorithm, to match the procedure of the study that originally proposed this approach. However, the random forest was outperformed by XGBoost [25], which was the method of choice for estimating non-linear relationships in all other feature-based modeling strategies. **Reads sensor-level signals but internally applies template-based source projection. ***ChronicPainNetworks, by definition, operates on network-level signals.

Crucially, increased model complexity did not translate to improved performance. Among the default configurations, best performance was achieved by the hypothesis-driven *ChronicPainNetworks* (R=0.11, R^2^=0.01), yielding moderate evidence for a positive prediction-observation correlation (BF=7.7). In comparison, the default configurations of data-driven models like *OTiS* (R=0.04) and *LaBraM* (R=0.00) did not detect resting-state EEG patterns predictive of chronic pain intensity.

When exploring the full design space, we identified specific configurations that outperformed the defaults for certain models. For *OTiS_EEG*, the optimized configuration achieved a correlation of R=0.08 (versus R=-0.02 by default). This improvement was driven by a parcel-level signal representation (100 Schaefer parcels), extended 20-second input segments, linear probing of the frozen backbone, and feature-level aggregation. However, despite these optimizations, the model failed to yield robust statistical evidence for a positive correlation with observed chronic pain intensity (BF=1.2). In sharp contrast, the optimized *FilterBankRiemann* model achieved the highest prediction-observation correlation across all modeling strategies (R=0.15, BF > 10^2^). This configuration relied on parcel-level signal representation (100 Schaefer parcels), providing strong evidence for an advantage over any other model considered (BF_FilterBankRiemann / BF_others > 10). Detailed performance metrics for all investigated configurations are presented in Supplementary Figures 1, 2, and 3.

### Age prediction benchmark

In contrast to the faint neural signatures of chronic pain, the positive control task of predicting chronological age from EEG confirmed that our modeling strategies were technically sound and capable of extracting biological information when it is robustly present, as summarized in Figure 6. All modeling strategies achieved strong performance in age prediction, with cross-validated correlations ranging from R=0.21 to R=0.53. The proportion of explained variance in age reached up to R^2^=0.27. As with the pain prediction task, rank-based prediction-observation correlations (Spearman’s rho) aligned closely with the parametric measures (Pearson’s R), confirming the robustness of the results against potential outliers.

**Figure 6:**
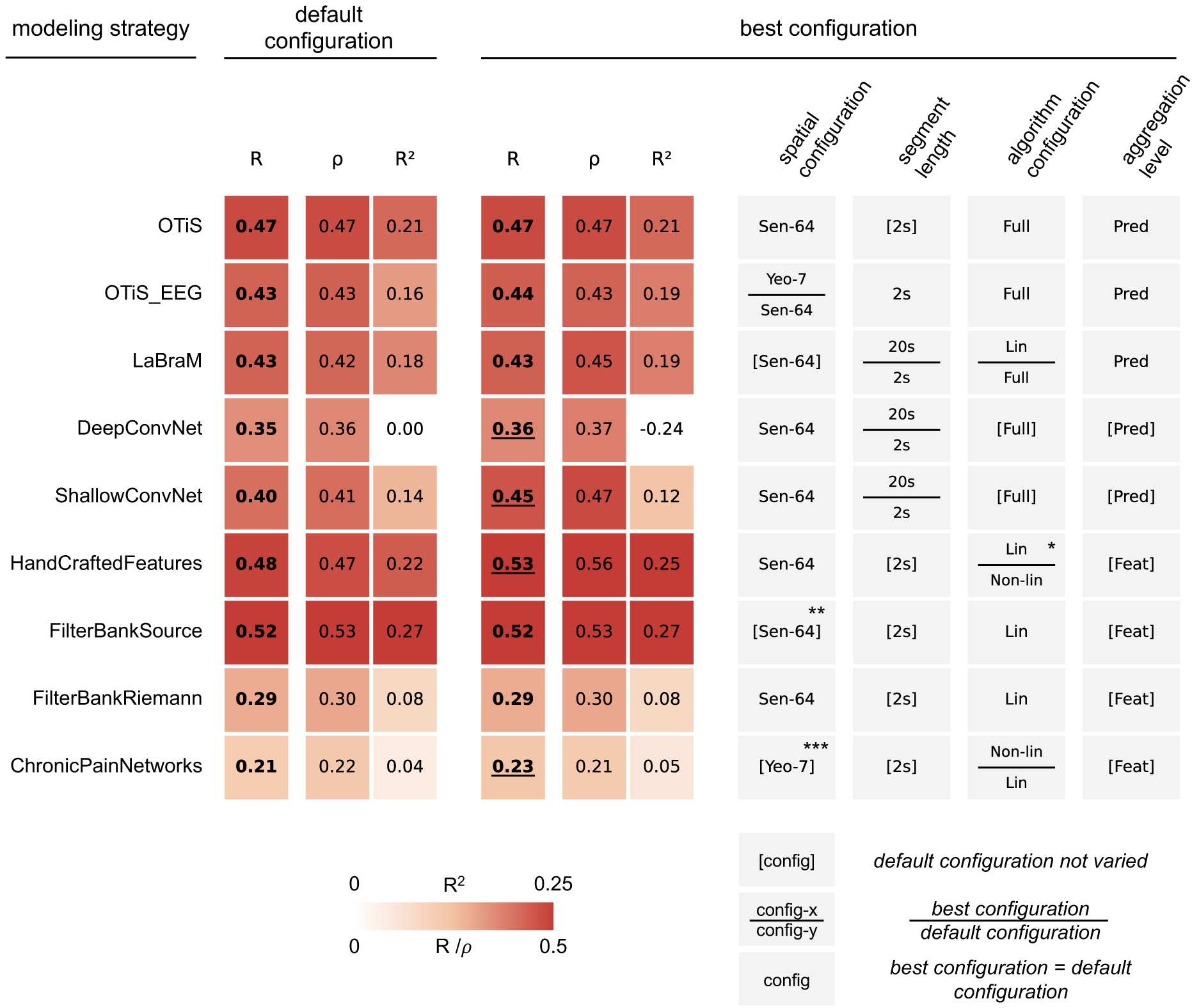
Age prediction performance of different modeling strategies. Cross-validated measures of association between predicted and observed age are reported (Pearson’s R; Spearman’s ρ; coefficient of determination R^2^). Scores in **bold** indicate at least moderate evidence for performance above chance (BF > 3). **(Left)** Default Configuration. This block summarizes the performance of each modeling strategy in its default configuration (see Figure 3). **(Right)** Best configuration. This block summarizes the performance of the single best configuration found for each modeling strategy, alongside its specific configuration details. <u>Underlined</u> scores indicate a BF ratio of best configuration versus default larger than 10. Brackets [] indicate configurations which were not varied for a given modeling strategy. Tiles divided by a horizontal bar indicate configurations for which the best configuration (top entry) differed from the default configuration (bottom entry). Sen-64: 64 sensor-level signals, Sch-100: 100 parcel-level signals, Yeo-7: 7 network-level signals, Lin: linear mapping from features to targets, Non-lin: non-linear mapping from features to targets, Full: full network training, Pred: prediction-level aggregation, Feat: feature-level aggregation. * The default configuration employed a non-linear mapping using a random forest algorithm, to match the procedure of the study that originally proposed this approach. **Reads sensor-level signals but internally applies template-based source projection. ***ChronicPainNetworks, by definition, operates on network-level signals.

Every modeling strategy yielded strong evidence for positive prediction-observation correlations in its default configuration (BF>10^5^). The best default model was *FilterBankSource* (R=0.52, R^2^=0.27, BF>10^40^), which showed strong evidence for superiority over the second-best default model, *HandCraftedFeatures* (R=0.48; BF_FilterBankSource / BF_HandCraftedFeatures > 10^6^).

The systematic exploration of configurations yielded further performance gains. For the convolutional neural networks (ConvNet), alternative configurations resulted in higher predictive accuracy than the default (BF_alternative / BF_default > 10). *ShallowConvNet* improved from R=0.40 to R=0.45, and *DeepConvNet* marginally from R=0.35 to R=0.36. Notably, for both models, best performance was achieved using the longer 20-second signal segments. The overall highest performance was achieved by the optimized HandCraftedFeatures (R=0.53, R^2^=0.25, BF = 10^43^; BF_HandCraftedFeatures / BF_others > 10^2^), which utilized a linear mapping of features to clinical target. Detailed performance metrics for all investigated configurations are presented in Supplementary Figures 4, 5, and 6.

The explained variances scores in age prediction were generally lower than those reported in a previous benchmarking study [13], which used different datasets and investigated only a subset of the modeling strategies considered in this study. This observation might be attributable to the rigorous harmonization of the target variable age through within-dataset z-scoring. To quantify this, we retrained the best model on non-harmonized targets, i.e. raw age values. This control experiment yielded markedly higher prediction-observation correlation and explained variance (R=0.60, R^2^=0.30, BF > 10^58^), approaching the performance levels reported previously. This confirms that the lower absolute scores in our main analysis reflect the stringent removal of confounding variance rather than model underperformance.

## Discussion

In this large-scale benchmarking study, we evaluated the potential and limitations of resting-state EEG as a source of biomarkers for inter-individual differences in chronic pain. By systematically benchmarking nine diverse modeling strategies, spanning conventional feature-based machine learning to state-of-the-art deep learning, across a multi-site dataset of 623 individuals with chronic pain, we draw two primary conclusions. First, cross-sectional resting-state EEG provides limited information about inter-individual differences in chronic pain intensity. Despite employing powerful data-driven strategies capable of extracting complex signal features, prediction performance remained modest (R=0.15) and fell short of thresholds required for clinically actionable biomarkers. Second, the technical capability of the modeling strategies to extract biological information from resting-state EEG was confirmed by the positive control task, as the models robustly decoded chronological age (R=0.53) from the same harmonized signals. This dissociation indicates that the limited performance in chronic pain intensity prediction is unlikely to reflect methodological insufficiency, but instead points towards intrinsic constraints in the information content of resting-state EEG regarding inter-individual differences in pain intensity.

The benchmarking of diverse modeling strategies yielded important insights into the nature of pain-related information in resting-state EEG. Contrary to the expectation that data-driven deep learning might uncover complex neural signatures of chronic pain beyond those identified by hypothesis-driven models, state-of-the-art Transformer architectures (*OTiS, LaBraM, OTiS_EEG*) and convolutional networks (*DeepConvNet, ShallowConvNet*) failed to outperform simpler machine learning baselines. Instead, the highest predictive accuracy was achieved by *FilterBankRiemann*, a conventional strategy utilizing features derived from signal covariances via Riemannian geometry. This reliance on functional connectivity suggests that any pain-related information in resting-state EEG is likely encoded in the statistical interactions between brain regions rather than in local signal features, replicating and modestly extending findings from prior work using the same dataset [1]. However, even the best model explained a very small amount of variance in pain ratings, confirming that EEG-based neural signatures of chronic pain intensity are currently insufficient for clinically actionable individual-level prediction.

The results of the age prediction task contrast to those of the pain prediction task and serve as a positive control. Across modeling strategies, chronological age was robustly decoded, confirming their sensitivity to biological information in resting-state EEG when such information is strongly expressed. As with pain prediction, conventional machine learning outperformed data-driven deep learning. However, the specific utility of modeling strategies differed significantly. Although pain prediction favored connectivity-based approaches, these approaches performed worst for age prediction. In contrast, the best performance for age prediction was achieved by *HandCraftedFeatures* and *FilterBankSource*, which rely on local signal features such as the spectral power across canonical frequency bands. This suggests that unlike the subtle neural signatures associated with chronic pain, chronological age is reflected in robust alterations of EEG power spectra that are already accessible at the sensor level. This dissociation demonstrates that successful age decoding does not imply sensitivity to neural signatures relevant for chronic pain.

An important finding of this study is the competitiveness and sometimes advantage of hypothesis-driven modeling over fully data-driven deep learning in the low-data regime of clinical EEG (N=623). Across both tasks, conventional machine learning, relying on *a priori defined* features and linear prediction heads, consistently matched or outperformed deep learning models, including pre-trained Transformers and convolutional neural networks. This pattern likely reflects the interplay between dataset size and model capacity. Although a sample size of N=623 is substantial for clinical EEG studies, it remains orders of magnitude smaller than the large-scale datasets typically required to train or fine-tune high-capacity Transformers. Interestingly, the general-purpose *OTiS* matched or even outperformed the specialized *LaBraM* and *OTiS_EEG*. Furthermore, given the inherently low signal-to-noise ratio in resting-state EEG, highly parameterized deep learning models may be particularly prone to overfitting in low-data regimes. In contrast, conventional models benefit from the strong inductive biases inherent in handcrafted, a priori defined features, which constrain the search space and improve robustness under low-data conditions.

The robust decoding of chronological age from the same resting-state EEG signals indicates that our processing pipeline, including harmonization, signal representation, feature extraction, and prediction, is effective in extracting biological information. Consequently, the limited performance in pain prediction is more likely to reflect biological or conceptual constraints which may include the following: First, resting-state EEG signals may contain little stable information about inter-individual differences. While it is not possible to exhaustively test all conceivable modeling approaches, the consistent failure of a broad and diverse set of methods to identify robust predictive patterns renders this interpretation plausible. Conversely, it seems unlikely that chronic pain-related EEG alterations, if consistently present across individuals, would be so specific as to evade detection by all evaluated approaches. Second, self-reported pain intensity may be an inappropriate target for biomarker development. Pain intensity captures only a single dimension of a complex, multidimensional disorder [26] and EEG correlates of chronic pain may be more strongly associated with other clinical features, such as affective or cognitive symptoms, which were not consistently available in the datasets analyzed here. Moreover, inter-individual differences in self-reported pain intensity may not only reflect differences in pain perception but also differences in scale usage, reporting style, or contextual interpretation not reflected in resting-state EEG signals. Third, measurement precision of resting-state EEG and clinical targets may have impaired reliability. EEG recordings were relatively short (< 2 minutes), and pain intensity was assessed using a single questionnaire item within a single session. Repeated measurements over time may be necessary to capture more stable pain-brain relationships across individuals [27]. Finally, the substantial clinical heterogeneity within our cohort of 623 individuals with chronic pain, which includes diverse pain etiologies, may have further limited the detectability of shared EEG-based biomarkers.

From an epistemological perspective, the biomarker discovery approach adopted here was nomothetic, aiming to identify population-level neural patterns that generalize across individuals based on cross-sectional data [28]. The findings suggest that applying deep learning to cross-sectional resting-state EEG is unlikely to yield actionable biomarkers of inter-individual differences in chronic pain intensity. However, this does not rule out the possibility that EEG activity might be helpful in developing other types of biomarkers [28–30]. Importantly, recent work on EEG-based decoding of acute experimental pain has demonstrated that models incorporating individual-specific components achieve higher explained variance and greater robustness than purely population-based approaches [31]. EEG may therefore be better suited for idiographic approaches that prioritize intra-individual variability over inter-individual differences. In practice, this would involve training individual-specific models to track fluctuations in pain over time. Such markers may be more detectable because they do not assume a shared neural representation across individuals. Extending this framework to chronic pain could improve mechanistic understanding and support clinical monitoring, including treatment-response tracking or use as surrogate endpoints in longitudinal trials.

In conclusion, this study represents the largest and most comprehensive benchmark of modeling strategies for EEG-based chronic pain biomarkers to date. Across this extensive benchmark, we found little support for using EEG-based neural patterns of inter-individual differences as clinically actionable biomarkers of chronic pain intensity. At the same time, the same modeling pipelines reliably decoded chronological age, demonstrating their in-principle capacity to extract meaningful individual characteristics from EEG signals. Together, these findings delineate current limits of cross-sectional EEG-based pain biomarkers and may motivate a shift towards more personalized, longitudinal approaches to brain-based chronic pain biomarker development.

## Methods

### Overview

In this study, we evaluated a broad range of data-driven modeling strategies for predicting chronic pain intensity from resting-state EEG data. Models were trained on EEG signal segments and produced pain intensity estimates as the primary target, while age prediction served as a secondary control target to verify the technical correctness of each implementation. Analyses were conducted on a large, multi-site dataset comprising resting-state EEG recordings collected across multiple international research centers.

### Datasets

In this study, we combined resting-state EEG data from individuals with chronic pain collected by multiple research groups worldwide. We acquired these data through a structured international data acquisition initiative. Specifically, we included EEG recordings previously collected by our own research group and invited research teams from Australia, Brazil, China, Denmark, Germany, Israel, Italy, New Zealand, Spain, and the US to collaborate and share their data. Candidate collaborators were identified based on a previously published systematic review of EEG studies in chronic pain populations, complemented by an updated PubMed search. This initiative yielded a comprehensive dataset of resting-state EEG recordings from more than 700 individuals. In the present study, data from 623 individuals satisfied our predefined quality criterion (≥6 clean 20s signal segments). The final sample comprised eight datasets originating from five different research groups. Detailed information on the data acquisition initiative and each dataset is available in the previous open-access publication [1]. Below, we briefly summarize the datasets included in the present analysis.

*Set_Munich1* (IRB number: 5493/12) is composed of multiple datasets that have previously been recorded in our research group to investigate brain dysfunction in people with chronic pain *[4, 32, 33]*. Throughout, EEG was recorded using a passive electrode EEG system with 64 channels (Easycap, Herrsching, Germany) and BrainAmp MR plus amplifier (Brain Products, Munich, Germany). Data from 117 individuals satisfied our quality criteria, including 73 with chronic back pain (CBP), 12 with chronic widespread pain (CWP), 20 with neuropathic pain (NP), and 12 with joint pain (JP).

*Set_Munich2* (IRB number: 6/22 S-KH) originates from a study examining the effects of pain medication on standard EEG features [34]. EEG was acquired using a 32-channel system with active dry electrodes (CGX-Quick32r, CGX-systems, San Diego, US). We here included data from 62 individuals: 18 with CBP, two with CWP, 17 with NP, 11 with JP, and 14 with pain of other etiologies (OTHER).

*Set_Brisbane* (IRB number: 2015000568) stems from a trial that evaluated the efficacy of several non-pharmacological, 8-week interventions for the treatment of CBP [35]. EEG was recorded using an ANT Neuro EEGO sports system (Medical Imaging Solutions GmbH, Berlin, Germany) with 64 active scalp electrodes (Waveguard cap). Baseline recordings from 64 individuals with CBP were included in the present study.

*Set_Otago1* (IRB number: 20/CEN/60) was collected as part of a study investigating the efficacy of infra-slow neurofeedback training as a treatment for chronic low back pain (CLBP) [36]. EEG was recorded using a 64-electrode system with SynAmps-RT amplifier (Compumeics-Neuroscan, Abbotford, Australia). For the analyses presented here, we used baseline data from 50 individuals with CLBP.

*Set_Otago2* (IRB numbers: 20/NTB/67, 2023 EXP 17953) was recorded as part of studies investigating the effects of neurofeedback and transcranial electrical stimulation on CBP. The dataset was collected by the same researchers, using equivalent recording conditions as used for the recording of *Set_Otago1*. Set_Otago2 eventually comprised data from 73 individuals with CBP.

*Set_Boulder* (IRB number: 16-0544) was recorded in the context of a larger study investigating the efficacy of pain reprocessing therapy for the treatment of CBP [37]. EEG was recorded using a 19-channel EEG system (Evoke system, evoke neuroscience, New York, USA). Here, we included data from 64 individuals with CBP.

*Set_Haifa* (IRB number: 0052-15-RMB) was recorded as part of the DOLORisk [38] initiative aiming to identify risk factors for the development and maintenance of neuropathic pain. EEG was recorded using a 64-channel EEG system with active electrodes (ActiCHamp, Brain Products, Munich, Germany). For the present analyses, we included data from 79 individuals with NP.

*Set_Seattle* (IRB number: 43605 G) stems from a study that investigated the relative effects of hypnotic cognitive therapy, standard cognitive therapy, hypnosis focused on pain reduction, and pain education in adults with a variety of chronic pain conditions [39]. EEG was recorded using a 128-channel hydrocell net connected to a GES 300 high-density EEG acquisition system (magnetism EGI, Eugene, USA). Here, we only included data from 114 individuals and from 116 EEG sensors, which were located in regions covered by the head model employed for source reconstruction. Of those, 44 had CBP, while the other 70 participants had chronic pain secondary to multiple sclerosis, spinal cord injury, or muscular dystrophy.

### Signal processing

#### Preprocessing

EEG recordings from all sites were preprocessed in a standardized manner using an automated pipeline proposed by [40] and adapted in [41] for the use of resting-state recordings. This pipeline integrates established functions from the Matlab-based EEGLAB toolbox [42]. It comprises following steps: downsampling to 250 Hz, line noise removal, bad channel rejection, re-referencing to the average reference, independent component analysis and automated rejection of independent components labelled by a machine learning classifier as “muscle” or “eye” components [43], and identification of artifact-contaminated segments. All preprocessing steps were executed using default parameter settings.

#### Segmentation

Preprocessed EEG signals were segmented into 20 seconds segments, or so called epochs, with 10 seconds overlaps. Segments containing artifact-marked data were excluded. To counteract potential biases from unbalanced segment number across participants, we retained exactly six clean epochs per individual and excluded individuals with fewer clean epochs. To evaluate the influence of segment length, we generated an additional dataset by subdividing each 20s segment into shorter 2s segments with 1s overlap. This was done such that no more than two 2s segments overlapped at any given time.

#### Sensor-level interpolation

Most standard variants of modeling strategies operated on input sequences representing sensor-level EEG signals. Because the EEG recordings differed across datasets in both the number and spatial placements of sensors, all signals were transformed into a unified layout using spatial interpolation. Specifically, signals were interpolated to a common 64-channel configuration based on the international 10-10 electrode placement system. To this end, we used spherical spline interpolation, implemented in FieldTrip’s sphsplint_from_FT.m function [44], with parameters set to order=4, degree=500, and λ=10^-5^.

#### Source reconstruction

Some variants of modeling strategies were trained on source-projected EEG signals representing brain activity in 100 brain parcels defined by the Schaefer atlas [45]. Source projection was performed using Linearly Constrained Minimum Variance (LCMV) beamformers [46] implemented in FieldTrip [44]. Broad-band (1-100 Hz), array-gain LCMV spatial filters were constructed based on a lead field and the broad-band covariance matrix. The lead field was computed by a boundary element approximation of the solution to the bioelectromagnetic forward problem, assuming a realistically shaped, three-shell head model and incorporating dataset-specific sensor geometry. To stabilize matrix inversion, Tikhonov regularization was applied at 5% of the mean sensor power. The fixed dipole orientation at each source location was selected to maximize the variance of the beamformer output. Finally, parcel-level time series were obtained by applying the broad-band LCMV operators to broadband, band-pass-filtered EEG data.

#### Spatial aggregation and representative time series

To reduce input dimensionality and leverage the potential informative value of a functional brain atlas, we spatially aggregated parcel-level signals within seven large-scale functional brain networks (also known as intrinsic brain networks) defined by the Yeo atlas [11]. This approach, which essentially extends a principal component decomposition by introducing additional orthogonality constraints, was shown in our previous study to be beneficial for the extraction of pain-related information from EEG signals. In short, it works by defining the representative signal of some brain structure (e.g., a functional brain network) as the one that maximises the explained variance across all signals associated with that brain structure while constraining the explained variance of signals associated with other brain structures to zero. Formally, consider network net_A_ for which we aim to estimate the representative signal 𝐫*^A^* ∈ ℝ*^n^* with 𝑛 time samples. Let 𝐀 ∈ ℝ*^kA×n^* denote the 𝑘*_A_* source-reconstructed signals associated with net_A_. In the proposed approach, the representative signal 𝐫*^A^* maximises the explained variance across the 𝑘*_A_* signals in 𝐀 while being orthogonal to a set of time series representing activity in the rest of the brain. This constrained optimization problem can be expressed as:

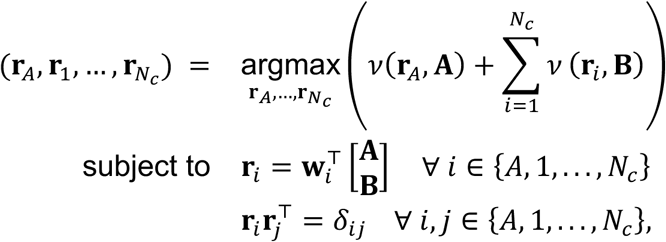

where 𝑁*_c_* denotes the number of components used to describe the activity outside of netA, 𝐫*_A_* is the signal representative of net_A_, and 𝜈(𝐫, 𝐗) is the fraction of the variance of matrix 𝐗 explained by vector 𝐫, i.e.,

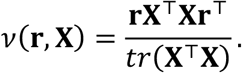

### General training and validation strategy

g Generalization performance of all models evaluated in this study, was estimated using 10-fold cross-validation (CV). This involved partitioning the entire dataset into ten equally-sized folds. In each iteration, nine folds were used for model training (training set) and the remaining fold was used for model testing (test set). This process was repeated ten times, such that each fold served as the test set exactly once. To avoid site-related biases, CV folds were stratified by dataset. Hyperparameter optimization followed a two-stage approach. In the first stage, during each outer CV-iteration, the training data were further subdivided into an inner training set and a validation set. Separate models were trained on the inner training set for each hyper parameter configuration. For deep learning models, which are computationally relatively expensive to train, we used a single split of the training set into an inner training and validation set. For traditional machine learning models, which are computationally relatively inexpensive to train, we employed nested CV. Here, the training set was partitioned into five folds, with four used for training and one for validation. Akin to the outer CV-loop, this was iterated such that each inner fold became the validation set exactly once. Validation set performance scores were then averaged across all inner CV folds. In the second stage, the model configuration achieving the highest validation-set performance score in the first training stage was retrained on the full training set and evaluated on the held-out test fold. An overview of explored hyperparameters for the individual modeling strategies is provided in supplementary Table S1.

To match the procedure used in our previous study [1], we removed age-related variability in pain intensity-prediction tasks where possible. Specifically, for all feature-based approaches, we fitted regression models predicting each individual features and pain intensity ratings as a function of age, and subsequently used these model’s residuals for further analyses.

### Harmonizing data across datasets

#### EEG-signal harmonization

To mitigate cross-site heterogeneity and enhance the detection of genuine associations between EEG signals and target variables, we harmonized EEG signals by adjusting signal power within each dataset. Specifically, signals were scaled by a dataset-specific normalization constant 𝑐*_norm_*(𝑑), where 𝑑 ∈ {1, . . ., 𝐷} and 𝐷 denotes the total number of datasets. This procedure preserved inter- and intra-individual variability in signal power, which, according to previous EEG-based pain studies [5], may carry important information about pain perception. Because the distribution of signal power across subjects follows a log-normal pattern, the constant 𝑐*_norm_*(𝑑) was chosen such that the mean logarithm of each subject’s total signal variance (i.e., “global power”) equaled 0 within each dataset. This condition leads to the expression

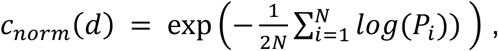

where 𝑁 represents the number of individuals within dataset 𝑑 and 𝑃*_i_* is the total signal variance of individual 𝑖. The total signal variance for each subject was computed as the mean variance across all channels:

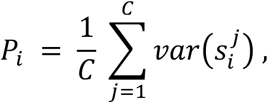

where 𝑠_𝑖_^𝑗^ is the signal in channel 𝑗 of subject 𝑖, and 𝐶 is the total number of channels. To prevent information leakage from test to training or validation sets, normalization constants were derived exclusively from the training and validation subsets, and computed separately for each CV-fold.

#### Target variable harmonization

To eliminate site-level variability and to minimize the risk of spurious site-related associations between EEG signals and Target variables, we standardized pain intensity and age within each dataset by computing z-scores. The mean and standard deviation used for z-transformation were estimated solely from the training and validation data within each CV-fold. This prevents information leakage from training and validation to test sets.

### Machine learning strategies

#### Overview

To comprehensively evaluate model performance across methodological paradigms, we implemented a diverse set of models spanning both deep learning and conventional machine learning approaches. These categories differ primarily in their approach to learn predictive representations, i.e., how the models extract informative structure from raw EEG data.

Conventional machine learning methods rely on predefined signal transformations to extract features, ideally designed based on prior domain knowledge. Their performance depends heavily on the quality and relevance of these handcrafted, manually extracted features. In contrast, deep learning models automatically learn feature representations directly from raw signals by jointly optimizing representation and prediction during training. This allows them to capture complex nonlinear dependencies, but this typically requires careful regularization and large datasets to prevent overfitting.

Within the deep learning models, we employed two architectural paradigms: convolutional neural networks (CNNs) and Transformers [12]. CNNs learn spatially or temporally local patterns through a sequence of filtering and pooling operations, making them particularly suited for tasks where relevant information is encoded in local signal motifs. Transformer-based architectures, in contrast, rely on self-attention mechanisms that allow the model to dynamically weight and integrate information across arbitrary temporal or spatial scales, enabling a more global signal analysis.

All Transformer-based models used in this study employed a two-step workflow including pre-training and subsequent fine-tuning. Pre-training refers to a self-supervised phase during which the model learns general-purpose representations from large unlabeled datasets. After pre-training, to predict the target of interest such as chronic pain intensity or age, the model is fine-tuned on a smaller, task-specific dataset annotated by experts. This two-step approach leverages transfer learning: representations acquired from broad diverse data can improve downstream performance even when labeled target data are scarce. In this work, pre-training of all Transformer models was performed following the masked data modeling paradigm: by reconstructing masked portions of the input signal, the model learns to extract meaningful and generalizable structures from the visible input. In contrast, CNNs and conventional strategies were trained directly on the target dataset without pre-training.

#### Transformer-based deep learning models (pre-training + fine-tuning)

**OTiS** [15] is a deep learning model based on a Transformer architecture [12], designed to learn general-purpose time series representations from large, heterogeneous datasets. It was pre-trained on a large, multi-domain corpus comprising both physiological signals (e.g., EEG, ECG) and non-physiological time series (e.g., weather, financial data) using a masked data modeling paradigm. OTiS introduces three key innovations to efficiently learn features from diverse data: First, a domain-specific tokeniser with learnable variate embeddings to capture the unique domain-dependent temporal and inter-channel relationships without requiring metadata. Second, a dual masking strategy to capture spatiotemporal structures and temporal causality. Third, a structure-aware reconstruction objective that integrates mean-squared error with a normalized cross-correlation term, encouraging the model to focus on signal shape and temporal alignment rather than just point-wise amplitude, effectively decoupling feature learning from modelling noise. Pre-trained model weights are publicly available at https://github.com/oetu/otis. For the present study, the model was re-pre-trained on the training subsets of the target dataset (independently for each fold of the outer cross-validation loop) and subsequently fine-tuned to predict chronic pain intensity or age. To preserve the generalized feature extraction capabilities of the encoder, re-pre-training was restricted to the variate embeddings, while all other model parameters were kept frozen, i.e. non-trainable.

**OTiS_EEG** [17] shares the same architecture and pre-training objective as OTiS but was pre-trained from scratch exclusively on large-scale resting-state EEG data (LEMON [47] and TDBrain [48]) to encourage domain specialization. This enables a fair evaluation of specialized EEG models by ruling out architectural, training, and scaling biases. Importantly, in our study, separate models were pre-trained for the three spatial input representations (see below) as these representations possess distinct physiological meanings. As pre-trained solely on EEG-derived signals, the different configurations of OTiS_EEG incorporate tokenisers whose variate embeddings specifically encode spatial relationships between neurophysiological signals at different levels of abstraction. For each outer cross-validation fold, the model was re-pre-trained on the corresponding training subset before being fine-tuned for pain intensity or age prediction.

**LaBraM** [16] is a Transformer-based deep learning model designed as a universal architecture for extracting EEG representations. It was pre-trained on a large corpus of over 2,500 hours of EEG data collected from 16 sites. These datasets cover heterogeneous recording configurations such as electrode numbers and sampling frequencies, and span a wide range of experimental paradigms including resting-state, motor imagery, emotion, and clinical recordings. The pre-training process involves a two-stage masked modeling strategy. In the first stage, a neural tokenizer was trained to convert raw EEG channel patches into discrete representations by predicting their spectral (Fourier-based) amplitude and phase components (instead of raw, sensor-level signals). This spectral prediction objective encourages the tokenizer to capture physiologically meaningful frequency patterns and improves training stability. In the second stage, the Transformer encoder was trained to reconstruct the original discrete neural tokens from a masked input, enabling the learning of robust contextual representations of neural dynamics. LaBraM introduces three key innovations: First, a vector-quantized codebook that provides a compact and semantically rich discrete representation of EEG patches. Second, a spatiotemporal embedding scheme combining learnable temporal and spatial embeddings that allows the model to accommodate EEG recordings with varying channel layouts and durations. Third, a symmetric masking strategy that enhances efficiency and representation diversity by jointly modeling masked and visible patch sets. Pre-trained model weights are available at https://github.com/935963004/LaBraM.

#### Convolutional Deep Learning models

**DeepConvNet** [18] is a deep convolutional neural network architecture designed for general-purpose EEG decoding, inspired by successful computer vision models. It comprises four convolution-max-pooling blocks and a final dense softmax output layer. To handle the high dimensionality of EEG channels, the first convolutional block employs a specialized two-step strategy: an initial temporal convolution learns frequency-specific filters, while a subsequent spatial filter learns to combine information across electrodes. This separation implicitly regularizes the model by forcing it to learn temporal dynamics and spatial patterns sequentially. Deeper layers utilize exponential linear units and standard max-pooling to extract increasingly abstract representations, allowing the model to learn diverse features from raw data without a priori selection.

**ShallowConvNet** [18] is a shallow convolutional neural network architecture specifically tailored to decode band-power features, inspired by the Filter Bank Common Spatial Pattern (FBCSP) algorithm (Ang et al., 2008). The architecture mimics the FBCSP pipeline through a sequence of trainable layers: a temporal convolution (analogous to bandpass filtering) and a spatial filter (analogous to CSP), followed by a squaring nonlinearity, mean pooling, and a logarithmic activation function. These final steps effectively compute the log-variance of the signal within specific frequency bands, a standard feature in EEG analysis. By integrating these operations into a single end-to-end differentiable model, ShallowConvNet allows for the joint optimization of the spectral and spatial filters alongside the classifier.

#### Conventional machine learning strategies

**HandCraftedFeatures** [20] represents a conventional machine learning strategy based on a diverse set of manually designed EEG signal summary statistics. These features are computed locally, i.e., per channel, and then concatenated across channels. They include basic time- and frequency-domain descriptors (e.g., standard deviation, kurtosis, skewness, and power ratios across canonical frequency bands), as well as information theoretic measures (e.g., estimates of spectral entropy, sample entropy, and temporal complexity). For prediction, this modeling strategy’s standard variant employs a random forest algorithm [49], chosen for its robustness to irrelevant features, capacity to model nonlinear relationships, and minimal sensitivity to hyperparameter tuning.

**FilterBankSource** [13, 21] is a conventional machine learning strategy based on source-reconstructed estimates of local brain activity. Source reconstruction is performed using a template-based minimum-norm estimation framework. Band-limited power values are then computed from the reconstructed source signals across 448 cortical regions and multiple canonical frequency bands, providing anatomically informed representations of neuronal activity for subsequent model training.

**FilterBankRiemann** [22, 23] is a conventional machine learning strategy deriving features from sensor-level covariance matrices computed within multiple frequency bands. These covariance features, which lie on a Riemannian manifold, are projected to a Euclidean tangent space to facilitate the use of conventional machine learning algorithms. As they capture statistical dependencies between signals, these features provide an implicit representation of interregional functional connectivity.

**ChronicPainNetworks** [1] is a conventional machine learning strategy based on source-reconstructed EEG features. Source reconstruction is performed using a Beamformer technique with a template-based head model. Source reconstructed signals are aggregated on the level of large-scale functional brain networks using a custom representative signals approach (see above). Amplitude-based connectivity values are then computed for all pairs of representative signals, providing functionally informed representations of neuronal communication for subsequent model training.

### Configuration of modeling strategies

To evaluate the ability of different modeling strategies to predict chronic pain intensity or age, we systematically investigated 72 distinct configurations per prediction task.

For most modeling strategies, the default spatial configuration consisted of input sequences representing 64 sensor-level signals. Alternatively, input sequences represented either 100 source-projected parcel-level (100 Schaefer parcels) signals or 7 aggregated network-level (7 Yeo networks) signals. By design, *LaBraM* operates exclusively on appropriately labeled sensor-level signals. Therefore, no spatial configuration variants were evaluated for this modeling strategy. For *ChronicPainNetworks*, the spatial configuration was inherently fixed to the 7 Yeo network-level signals. *FilterBankSource* nominally operates on sensor-level data but internally transforms these signals into a parcel-based source-level representation. Consequently, no alternative spatial configurations were explored for this approach. Although the theoretical formulation of FilterBankRiemann implicitly incorporates source-projection-like signal unmixing, we additionally evaluated this method using 100 parcel-based source-level signals because this configuration was technically feasible and required minimal additional implementation effort. We did not evaluate it using the 7 network-level signals because these representations are, by construction, largely orthogonalized, making covariance estimation largely uninformative.

The default temporal length of input sequences across all modeling strategies was 2-seconds. In addition, a sequence length of 20-seconds was evaluated for all deep learning-based strategies, with the exception of *OTiS*, for which 20-seconds sequences exceeded the available hardware memory. For conventional modeling strategies, which rely on features that are largely invariant to the duration of the signals from which they are computed, input sequence length was not varied.

For deep learning-based strategies, the default algorithmic configuration involved full network fine-tuning. For Transformer-based models, we additionally evaluated an embeddings-as-features paradigm, in which learned embedding vectors were mapped to the target variable using either linear or non-linear models. Linear mapping was implemented using principal component regression. This procedure involved computing a principal component decomposition of the embedding vectors across individuals in the training set, followed by selection of a subset of components as predictors in a standard least-squares regression model. The optimal number of retained components was determined using nested cross-validation, as described above. Non-linear mapping was implemented similarly but replacing standard least squares regression with XGBoost [25], a tree-based ensemble method, with hyperparameters similarly optimized via nested cross-validation. For conventional modeling strategies, we evaluated the same linear and non-linear feature-to-target mappings.

Finally, for Transformer-based models, we varied the strategy used to aggregate information across multiple signal segments within individuals. Specifically, we either averaged the embedding features across all segments for each individual and generated a single prediction from the aggregated representation, or generated one prediction per segment and subsequently averaged these predictions at the individual level.

Figures 2 and 3 illustrate all modeling strategies and their corresponding variants. Figures S1 and S2 provide a comprehensive overview of all modeling strategies evaluated in this study.

## Data Availability

Data produced in the present study are available upon reasonable request to the authors

## Acknowledgements

The study was supported by the Deutsche Forschungsgemeinschaft (SFB1158, PL321/14-1, PL321/16-1) and the Technical University of Munich (TUM Innovation Network *Neurotech*).

## Author contributions

Felix S. Bott: Conceptualization, Methodology, Software, Formal analysis, Investigation, Data curation, Writing – original draft, Writing – review & editing, Visualization

Özgün Turgut: Conceptualization, Methodology, Software, Writing – review & editing, Visualization

Paul Theo Zebhauser: Investigation, Resources, Writing – review & editing

Divya B. Adhia: Investigation, Resources, Writing – review & editing

Yoni K. Ashar: Investigation, Resources, Writing – review & editing

Melissa A. Day: Investigation, Resources, Writing – review & editing

Yelena Granovsky: Investigation, Resources, Writing – review & editing

David Yarnitsky: Investigation, Resources, Writing – review & editing

Mark P. Jensen: Investigation, Resources, Writing – review & editing

Tor D. Wager: Investigation, Resources, Writing – review & editing

Daniel Rueckert: Methodology, Writing – review & editing, Project administration, Funding acquisition

Markus Ploner: Conceptualization, Resources, Writing – original draft, Writing – review & editing, Visualization, Supervision, Project administration, Funding acquisition

## Materials availability

This study did not generate new unique reagents.

## Data and code availability

The EEG and meta data for the datasets Set_Brisbane, Set_Otago, Set_Boulder, Set_Haifa, and Set_Seattle are not deposited in a public repository due to formal data sharing agreements with the collaborating research institutions. The EEG and meta data of dataset Set_Munich1 are publicly available at https://osf.io/srpbg/. The EEG and meta data of dataset Set_Munich2 cannot yet be deposited in a public repository as they are part of an ongoing study. Public access to Set_Munich2 will be provided upon completion and publication of the study. A corresponding link will be provided at https://osf.io/9j3xh/. We will provide the code via the following download link upon acceptance of the manuscript for publication or request by the editor: https://drive.google.com/drive/folders/1dal5PHh3bSeqeoou8sZWRKzWgFEHv01p?usp=share_link. Any additional information required to reanalyze the data reported in this paper is available from the lead contact upon request.

## Declaration of interests

The authors declare no competing interests.

## Declaration of generative AI and AI-assisted technologies in the writing process

During the preparation of this work the authors used chatGPT 4o in order to improve the readability and language of this manuscript. After using this tool, the authors reviewed and edited the content as needed and take full responsibility for the content of the published article.

## Supplementary materials

**Figure S1:**
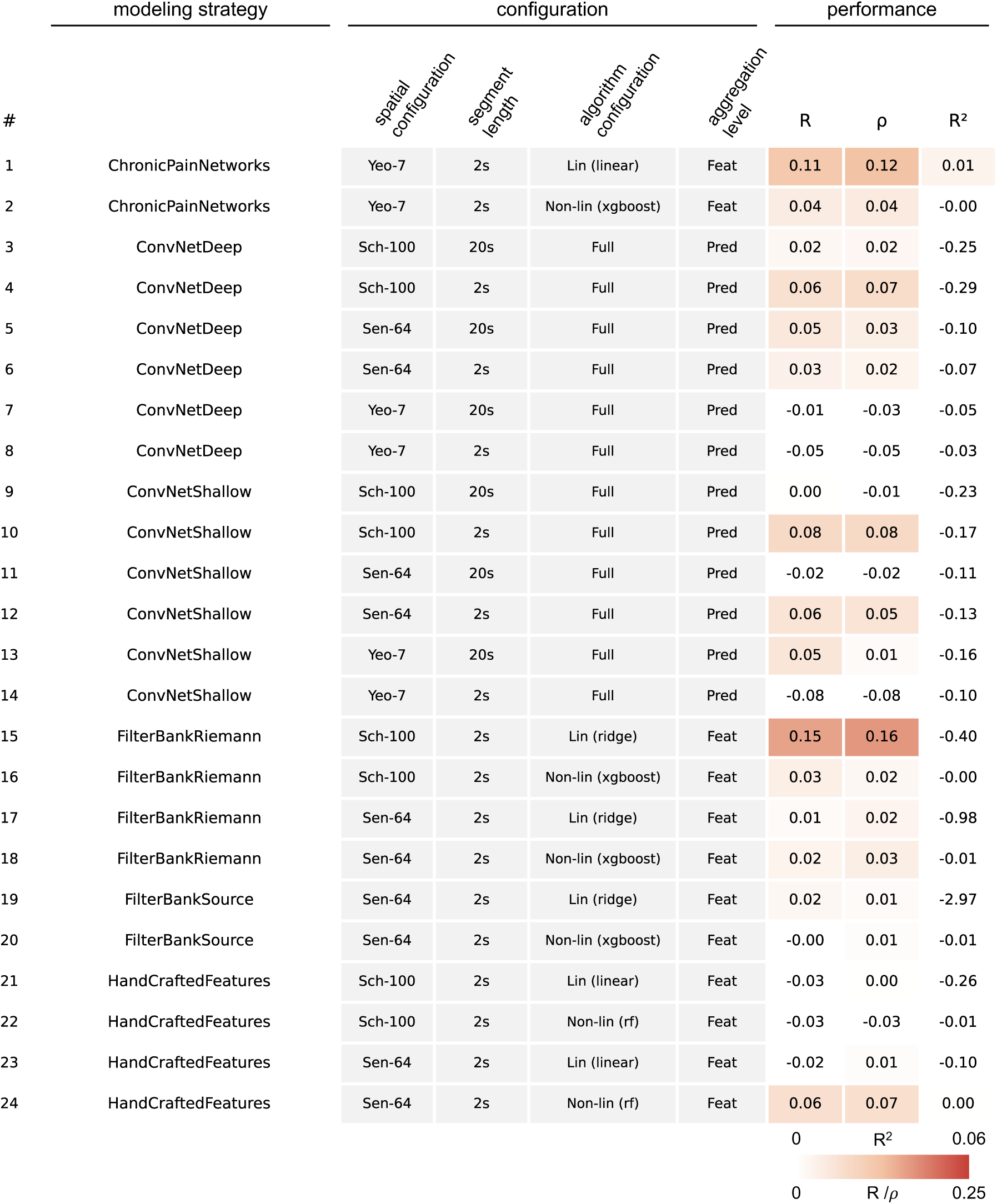
Overview of investigated pain prediction models and their configuration variants (spatial configuration, segment length, algorithm, and aggregation level), together with 10-fold cross-validated performance metrics. This figure shows models 1–24 out of a total of 72.

**Figure S2:**
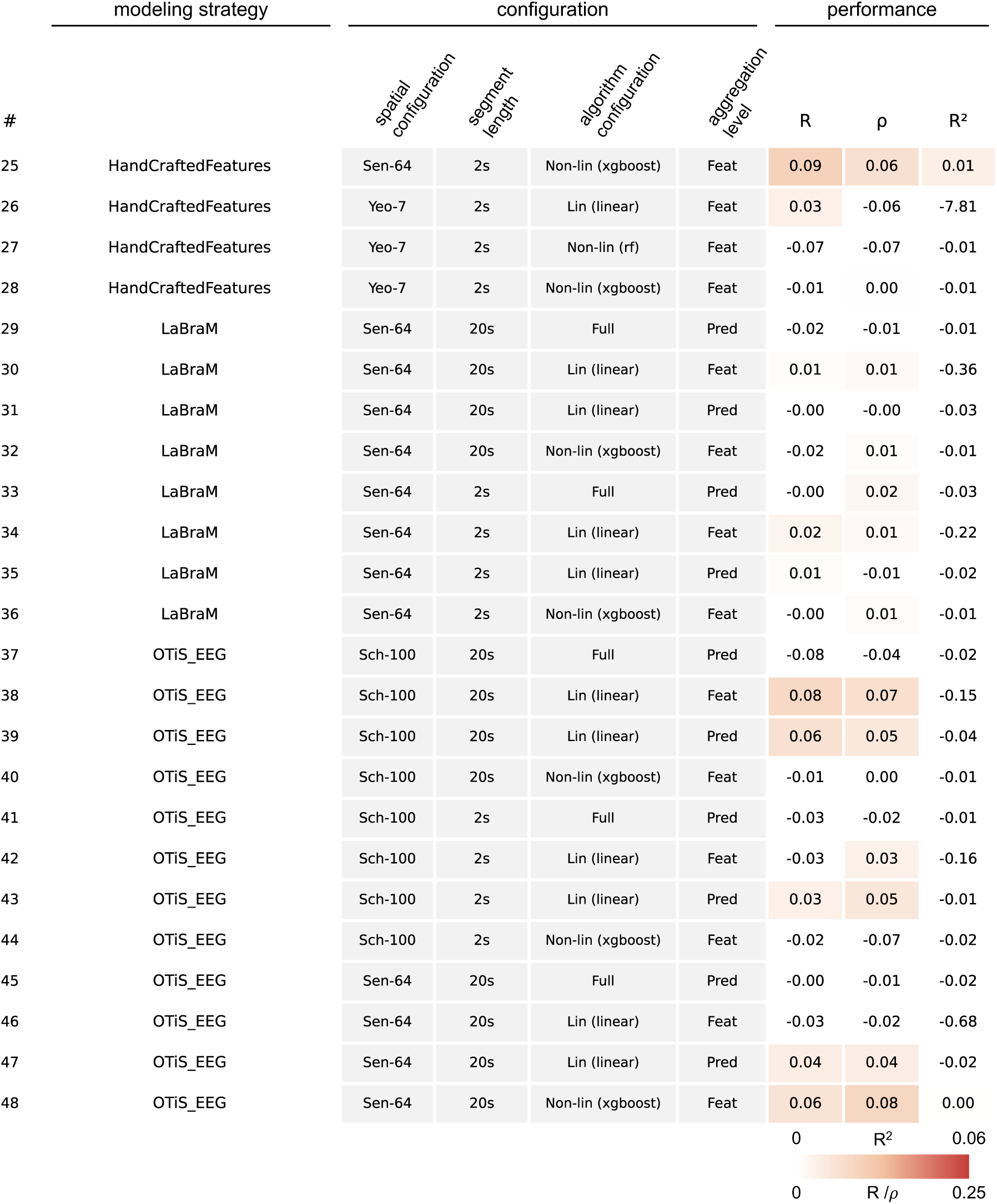
Overview of investigated pain prediction models and their configuration variants (spatial configuration, segment length, algorithm, and aggregation level), together with 10-fold cross-validated performance metrics. This figure shows models 25–48 out of a total of 72.

**Figure S3:**
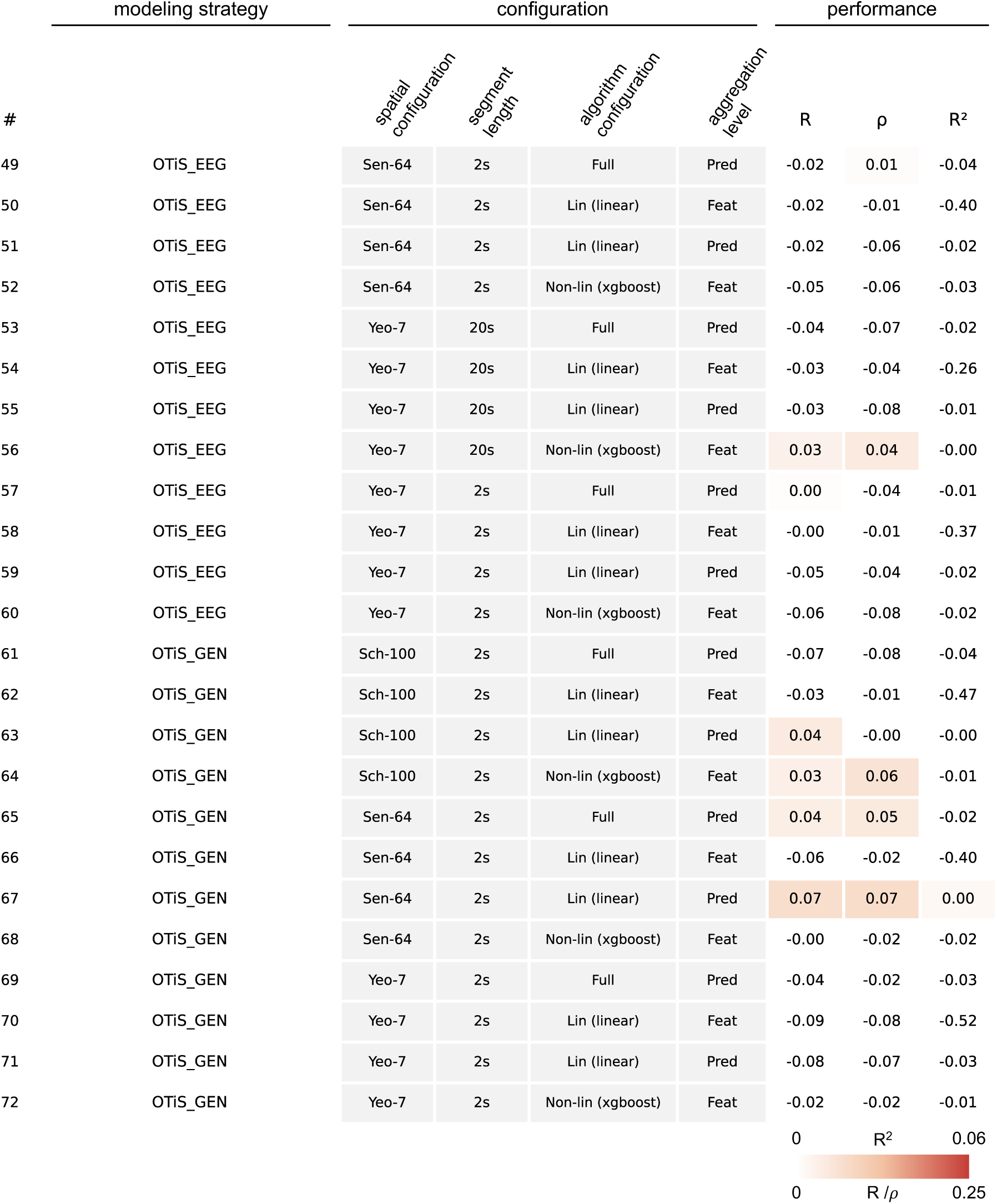
Overview of investigated pain prediction models and their configuration variants (spatial configuration, segment length, algorithm, and aggregation level), together with 10-fold cross-validated performance metrics. This figure shows models 49–72 out of a total of 72.

**Figure S4:**
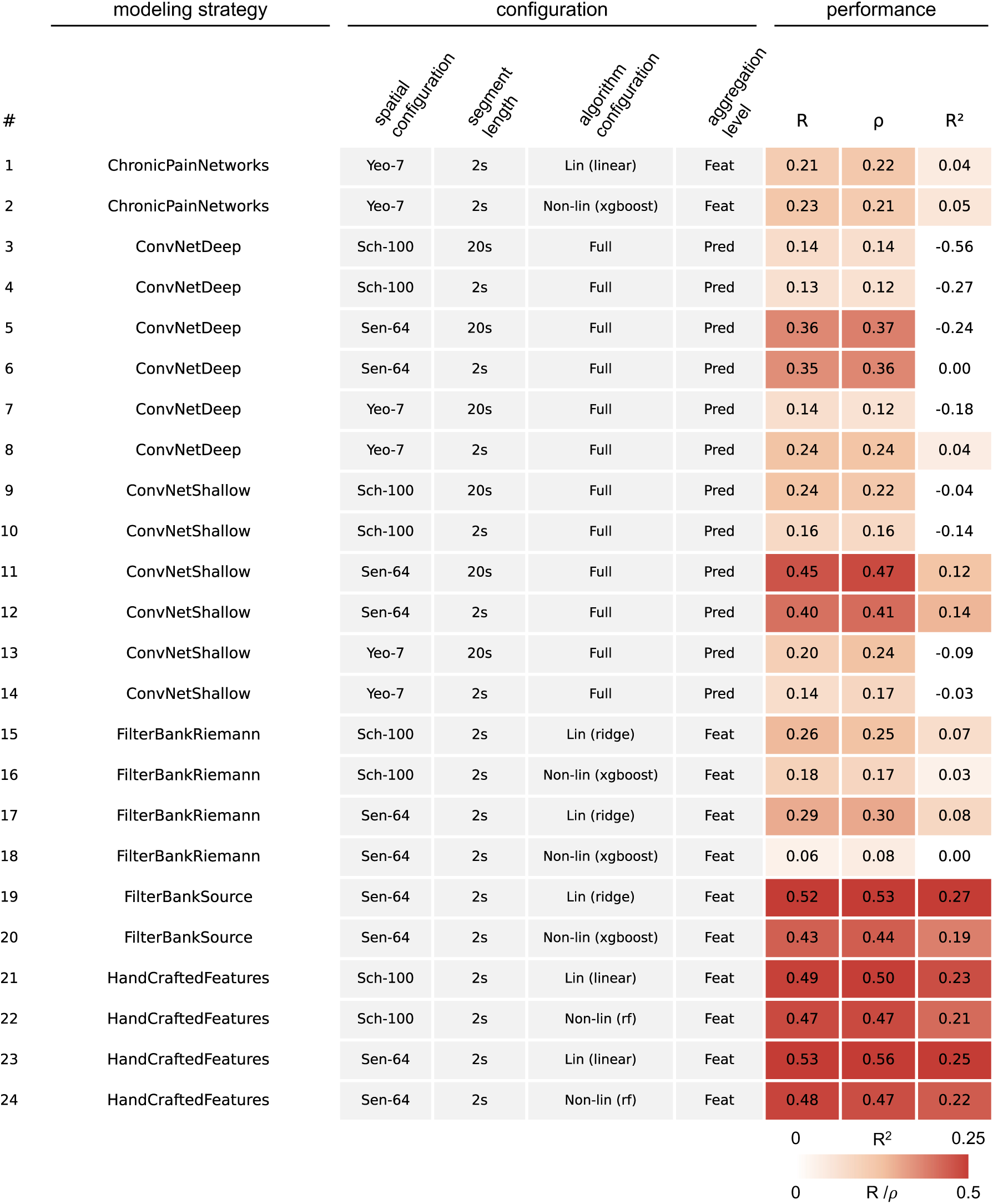
Overview of investigated age prediction models and their configuration variants (spatial configuration, segment length, algorithm, and aggregation level), together with 10-fold cross-validated performance metrics. This figure shows models 1–24 out of a total of 72.

**Figure S5:**
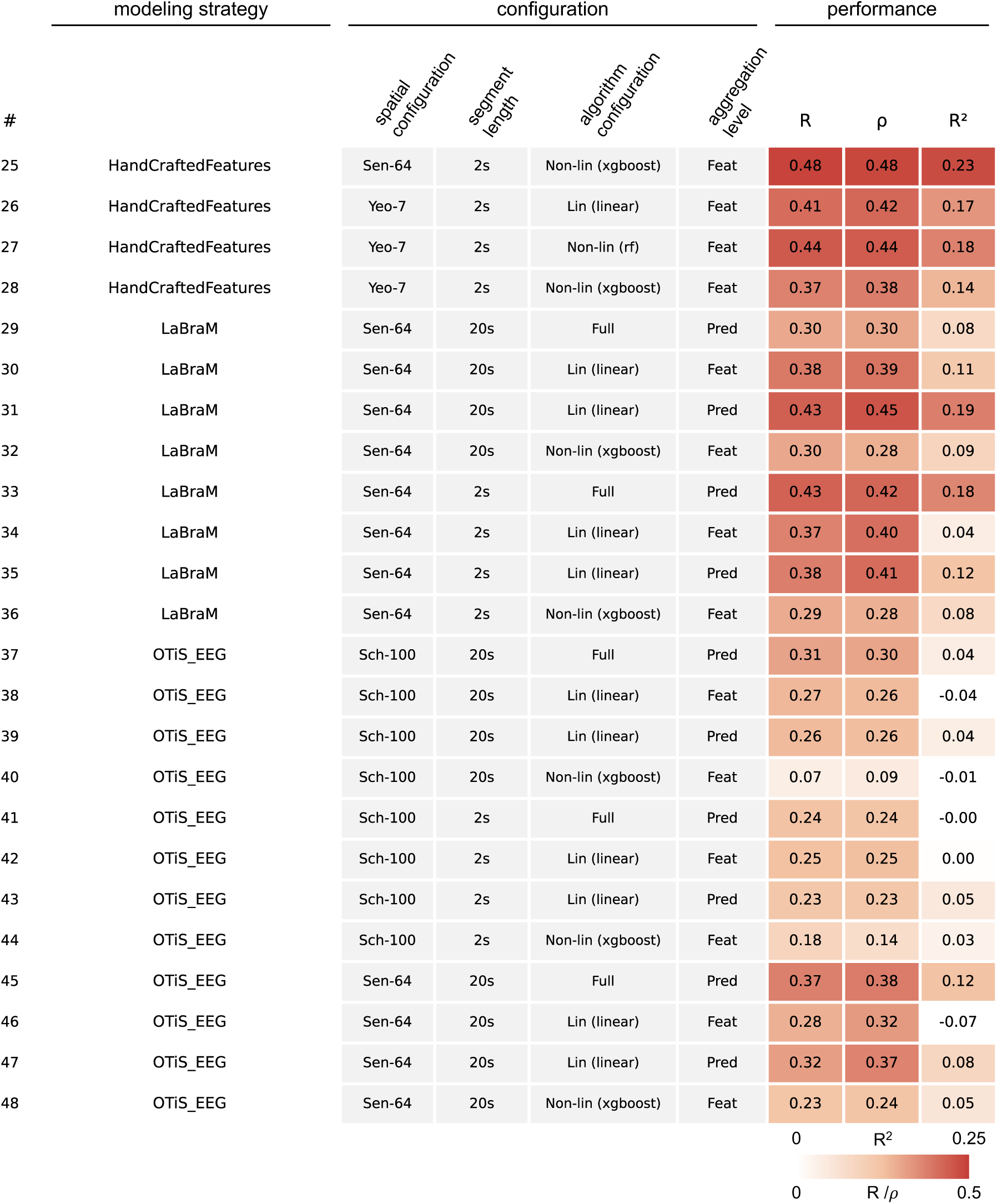
Overview of investigated age prediction models and their configuration variants (spatial configuration, segment length, algorithm, and aggregation level), together with 10-fold cross-validated performance metrics. This figure shows models 25–48 out of a total of 72.

**Figure S6:**
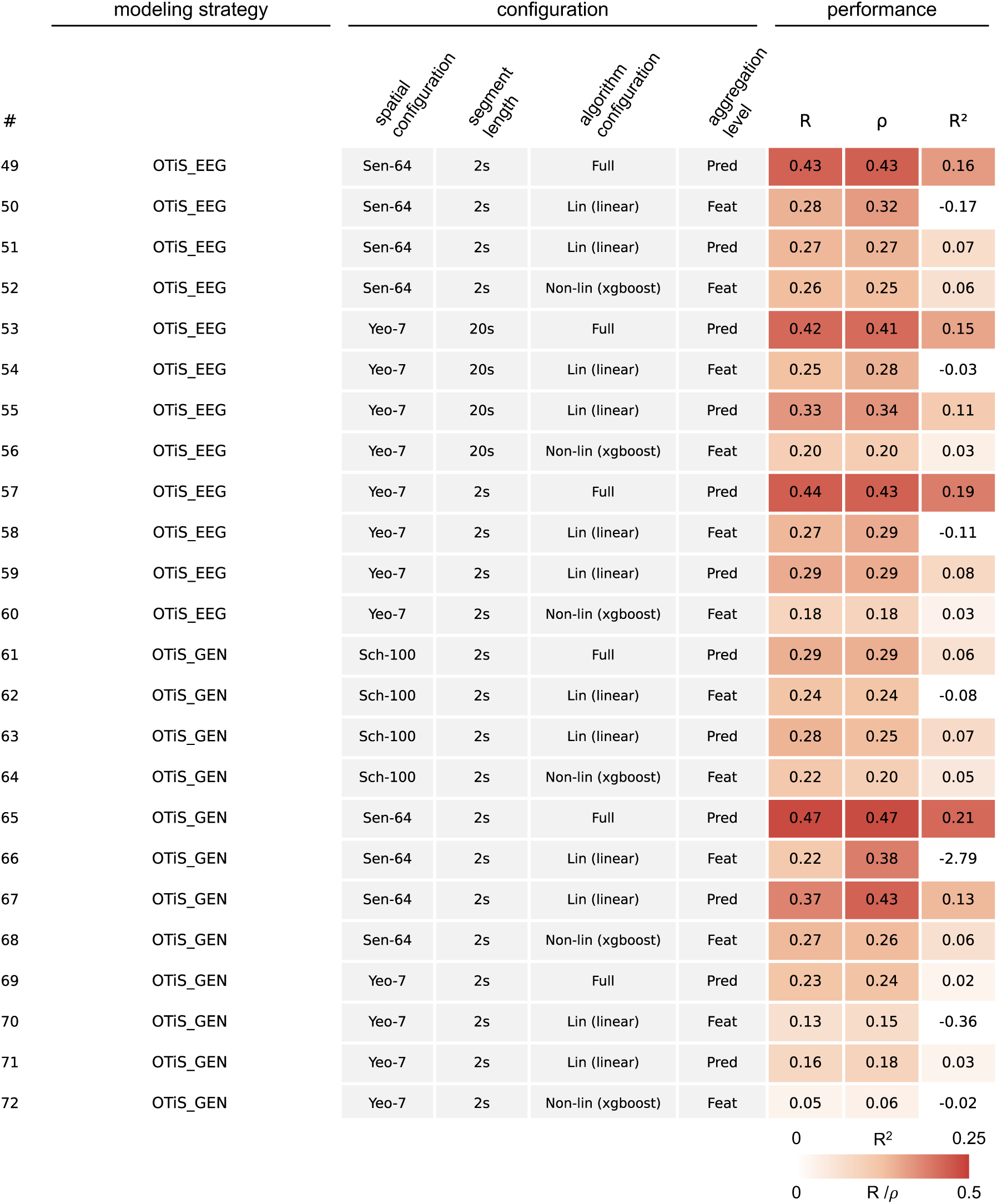
Overview of investigated age prediction models and their configuration variants (spatial configuration, segment length, algorithm, and aggregation level), together with 10-fold cross-validated performance metrics. This figure shows models 49–72 out of a total of 72.

**Table T1:**
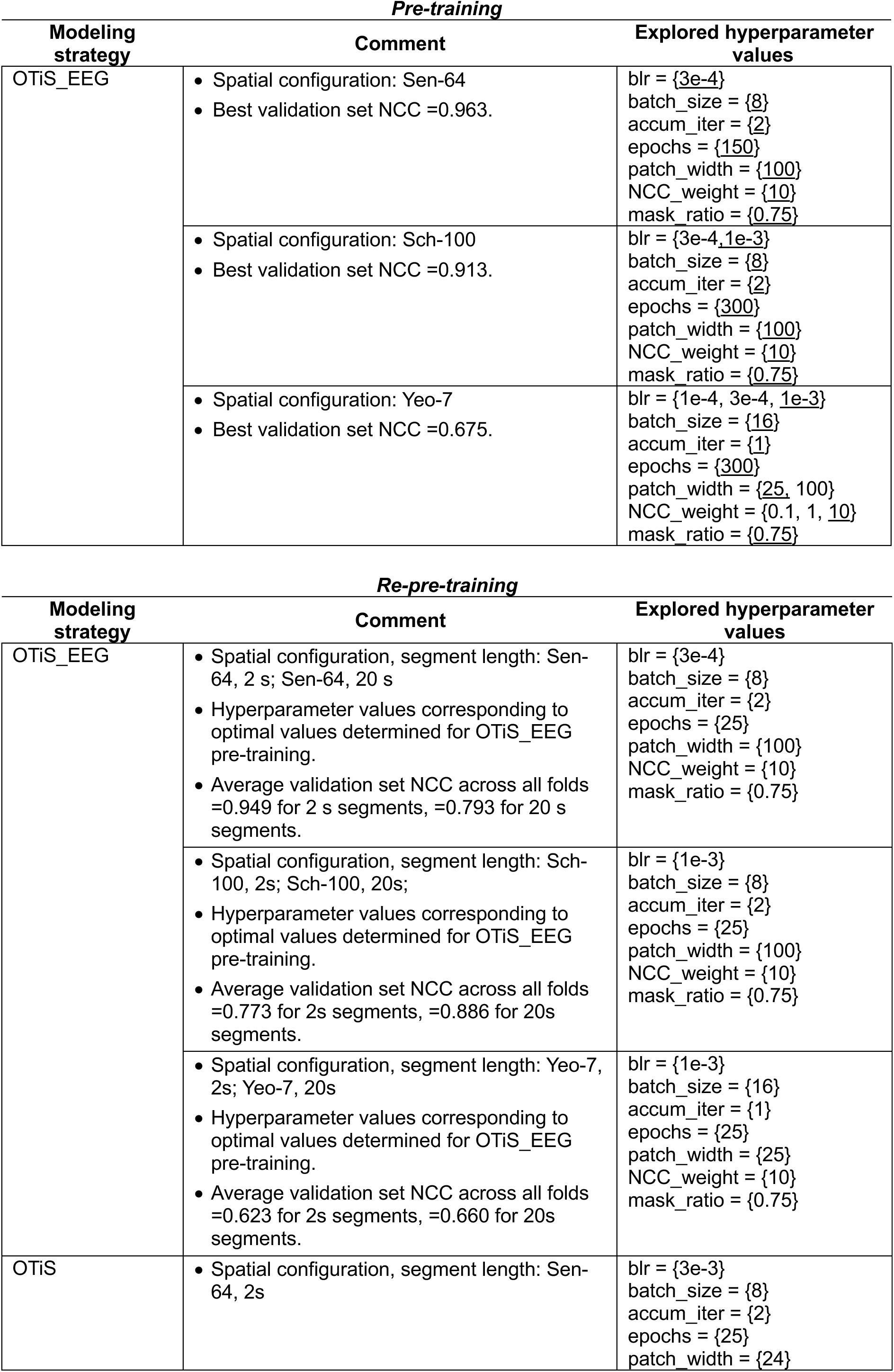
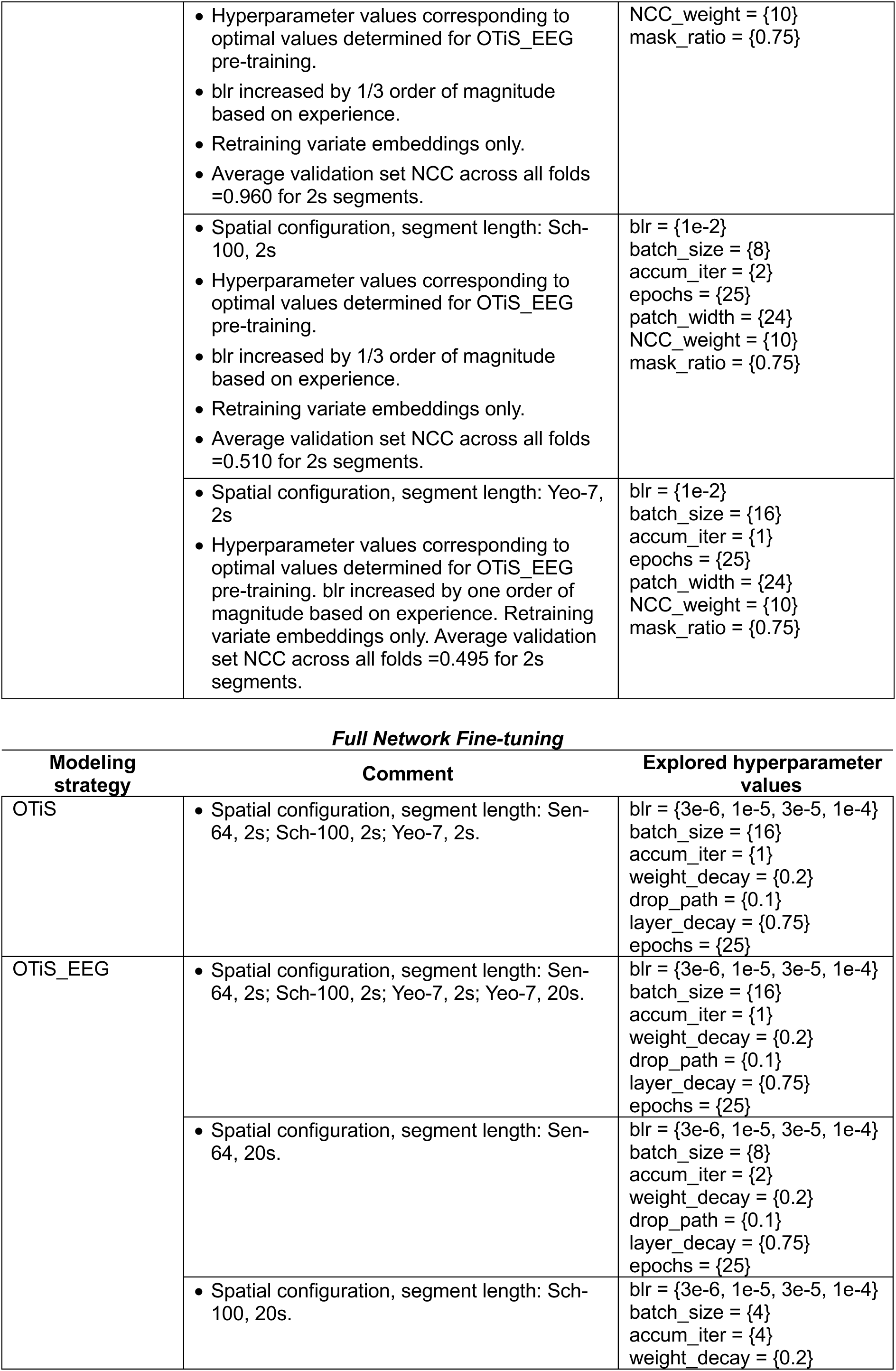
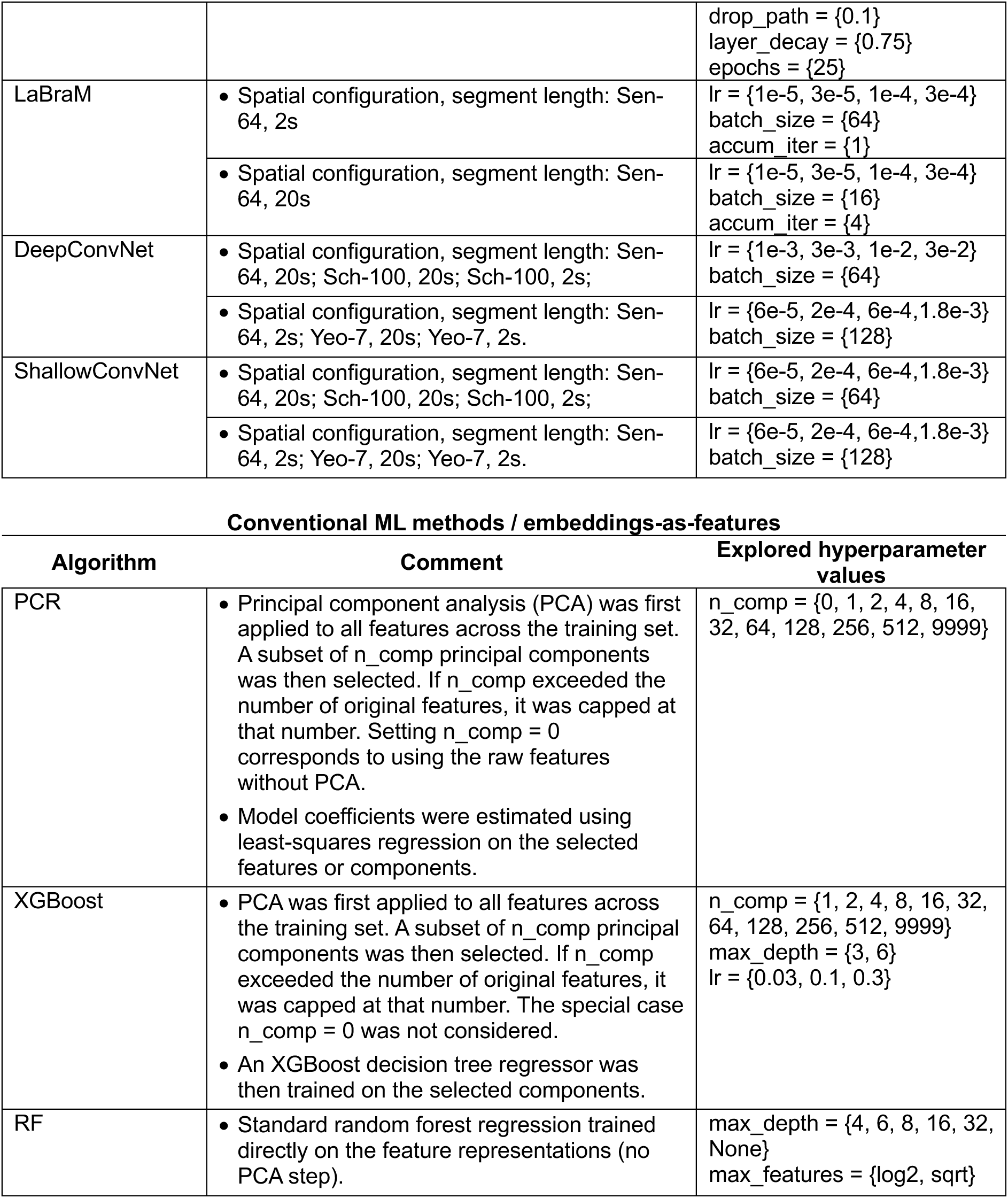
Overview of explored hyper parameters for each method.

**Pre-training**
| Modeling strategy | Comment | Explored hyperparameter values |
| --- | --- | --- |
| OTiS_EEG | Spatial configuration: Sen-64<br>Best validation set NCC =0.963. | blr = {3e-4}<br>batch_size = {8}<br>accum_iter = {2}<br>epochs = {150}<br>patch_width = {100}<br>NCC_weight = {10}<br>mask_ratio = {0.75} |
|  | Spatial configuration: Sch-100<br>Best validation set NCC =0.913. | blr = {3e-4, 1e-3}<br>batch_size = {8}<br>accum_iter = {2}<br>epochs = {300}<br>patch_width = {100}<br>NCC_weight = {10}<br>mask_ratio = {0.75} |
|  | Spatial configuration: Yeo-7<br>Best validation set NCC =0.675. | blr = {1e-4, 3e-4, 1e-3}<br>batch_size = {16}<br>accum_iter = {1}<br>epochs = {300}<br>patch_width = {25, 100}<br>NCC_weight = {0.1, 1, 10}<br>mask_ratio = {0.75} |

**Re-pre-training**
| Modeling strategy | Comment | Explored hyperparameter values |
| --- | --- | --- |
| OTiS_EEG | Spatial configuration, segment length: Sen-64, 2 s; Sen-64, 20 s<br>Hyperparameter values corresponding to optimal values determined for OTiS_EEG pre-training.<br>Average validation set NCC across all folds =0.949 for 2 s segments, =0.793 for 20 s segments. | blr = {3e-4}<br>batch_size = {8}<br>accum_iter = {2}<br>epochs = {25}<br>patch_width = {100}<br>NCC_weight = {10}<br>mask_ratio = {0.75} |
|  | Spatial configuration, segment length: Sch-100, 2s; Sch-100, 20s;<br>Hyperparameter values corresponding to optimal values determined for OTiS_EEG pre-training.<br>Average validation set NCC across all folds =0.773 for 2s segments, =0.886 for 20s segments. | blr = {1e-3}<br>batch_size = {8}<br>accum_iter = {2}<br>epochs = {25}<br>patch_width = {100}<br>NCC_weight = {10}<br>mask_ratio = {0.75} |
|  | Spatial configuration, segment length: Yeo-7, 2s; Yeo-7, 20s<br>Hyperparameter values corresponding to optimal values determined for OTiS_EEG pre-training.<br>Average validation set NCC across all folds =0.623 for 2s segments, =0.660 for 20s segments. | blr = {1e-3}<br>batch_size = {16}<br>accum_iter = {1}<br>epochs = {25}<br>patch_width = {25}<br>NCC_weight = {10}<br>mask_ratio = {0.75} |
| OTiS | Spatial configuration, segment length: Sen-64, 2s | blr = {3e-3}<br>batch_size = {8}<br>accum_iter = {2}<br>epochs = {25}<br>patch_width = {24} |
|  | Hyperparameter values corresponding to optimal values determined for OTiS_EEG pre-training.<br>blr increased by 1/3 order of magnitude based on experience.<br>Retraining variate embeddings only.<br>Average validation set NCC across all folds =0.960 for 2s segments. | NCC_weight = {10}<br>mask_ratio = {0.75} |
|  | Spatial configuration, segment length: Sch-100, 2s<br>Hyperparameter values corresponding to optimal values determined for OTiS_EEG pre-training.<br>blr increased by 1/3 order of magnitude based on experience.<br>Retraining variate embeddings only.<br>Average validation set NCC across all folds =0.510 for 2s segments. | blr = {1e-2}<br>batch_size = {8}<br>accum_iter = {2}<br>epochs = {25}<br>patch_width = {24}<br>NCC_weight = {10}<br>mask_ratio = {0.75} |
|  | Spatial configuration, segment length: Yeo-7, 2s<br>Hyperparameter values corresponding to optimal values determined for OTiS_EEG pre-training. blr increased by one order of magnitude based on experience. Retraining variate embeddings only. Average validation set NCC across all folds =0.495 for 2s segments. | blr = {1e-2}<br>batch_size = {16}<br>accum_iter = {1}<br>epochs = {25}<br>patch_width = {24}<br>NCC_weight = {10}<br>mask_ratio = {0.75} |

**Full Network Fine-tuning**
| **Modeling strategy** | **Comment** | **Explored hyperparameter values** |
| --- | --- | --- |
| OTiS | Spatial configuration, segment length: Sen-64, 2s; Sch-100, 2s; Yeo-7, 2s. | blr = {3e-6, 1e-5, 3e-5, 1e-4}<br>batch_size = {16}<br>accum_iter = {1}<br>weight_decay = {0.2}<br>drop_path = {0.1}<br>layer_decay = {0.75}<br>epochs = {25} |
| OTiS_EEG | Spatial configuration, segment length: Sen-64, 2s; Sch-100, 2s; Yeo-7, 2s; Yeo-7, 20s. | blr = {3e-6, 1e-5, 3e-5, 1e-4}<br>batch_size = {16}<br>accum_iter = {1}<br>weight_decay = {0.2}<br>drop_path = {0.1}<br>layer_decay = {0.75}<br>epochs = {25} |
|  | Spatial configuration, segment length: Sen-64, 20s. | blr = {3e-6, 1e-5, 3e-5, 1e-4}<br>batch_size = {8}<br>accum_iter = {2}<br>weight_decay = {0.2}<br>drop_path = {0.1}<br>layer_decay = {0.75}<br>epochs = {25} |
|  | Spatial configuration, segment length: Sch-100, 20s. | blr = {3e-6, 1e-5, 3e-5, 1e-4}<br>batch_size = {4}<br>accum_iter = {4}<br>weight_decay = {0.2} |
|  |  | drop_path = {0.1}<br>layer_decay = {0.75}<br>epochs = {25} |
| LaBraM | • Spatial configuration, segment length: Sen-64, 2s | lr = {1e-5, 3e-5, 1e-4, 3e-4}<br>batch_size = {64}<br>accum_iter = {1} |
|  | • Spatial configuration, segment length: Sen-64, 20s | lr = {1e-5, 3e-5, 1e-4, 3e-4}<br>batch_size = {16}<br>accum_iter = {4} |
| DeepConvNet | • Spatial configuration, segment length: Sen-64, 20s; Sch-100, 20s; Sch-100, 2s; | lr = {1e-3, 3e-3, 1e-2, 3e-2}<br>batch_size = {64} |
|  | • Spatial configuration, segment length: Sen-64, 2s; Yeo-7, 20s; Yeo-7, 2s. | lr = {6e-5, 2e-4, 6e-4, 1.8e-3}<br>batch_size = {128} |
| ShallowConvNet | • Spatial configuration, segment length: Sen-64, 20s; Sch-100, 20s; Sch-100, 2s; | lr = {6e-5, 2e-4, 6e-4, 1.8e-3}<br>batch_size = {64} |
|  | • Spatial configuration, segment length: Sen-64, 2s; Yeo-7, 20s; Yeo-7, 2s. | lr = {6e-5, 2e-4, 6e-4, 1.8e-3}<br>batch_size = {128} |

**Table T1:** Overview of explored hyper parameters for each method.
| Algorithm | Comment | Explored hyperparameter values |
| --- | --- | --- |
| PCR | Principal component analysis (PCA) was first applied to all features across the training set. A subset of n_comp principal components was then selected. If n_comp exceeded the number of original features, it was capped at that number. Setting n_comp = 0 corresponds to using the raw features without PCA.<br>Model coefficients were estimated using least-squares regression on the selected features or components. | n_comp = {0, 1, 2, 4, 8, 16, 32, 64, 128, 256, 512, 9999} |
| XGBoost | PCA was first applied to all features across the training set. A subset of n_comp principal components was then selected. If n_comp exceeded the number of original features, it was capped at that number. The special case n_comp = 0 was not considered.<br>An XGBoost decision tree regressor was then trained on the selected components. | n_comp = {1, 2, 4, 8, 16, 32, 64, 128, 256, 512, 9999}<br>max_depth = {3, 6}<br>lr = {0.03, 0.1, 0.3} |
| RF | Standard random forest regression trained directly on the feature representations (no PCA step). | max_depth = {4, 6, 8, 16, 32, None}<br>max_features = {log2, sqrt} |

## Notes

### Competing Interest Statement

The authors have declared no competing interest.

### Author Declarations

This study involved secondary analysis of de-identified, anonymised data obtained from controlled-access repositories. No new data were collected, and there was no interaction with or intervention involving human participants. As such, this analysis does not constitute human subjects research and did not require additional ethical approval. All original studies from which data were derived had received appropriate ethical approval: Set_Munich1: IRB of Technical University of Munich gave ethical approval for this work (IRB-number: 5493/12) Set_Munich2: IRB of Technical University of Munich gave ethical approval for this work (IRB number: 6/22 S-KH) Set_: IRB of Technical University of Munich gave ethical approval for this work (IRB-number: 5493/12) Set_Brisbane: IRB of University of Queensland gave ethical approval for this work (IRB number: 6/22 S-KH) Set_Otago1: IRB of University of Otago gave ethical approval for this work (IRB number: 20/CEN/60) Set_Otago2: IRB of University of Otago gave ethical approval for this work (IRB numbers: 20/NTB/67, 2023 EXP 17953) Set_Boulder: IRB of Dartmouth College gave ethical approval for this work (IRB-number: 5493/12) Set_Haifa: IRB of Technion Israel Institute of Technology gave ethical approval for this work (IRB number: 6/22 S-KH) Set_Seattle: IRB of University of Washington gave ethical approval for this work (IRB-number: 5493/12)

